# Distinct immune responses in patients infected with influenza or SARS-CoV-2, and in COVID-19 survivors, characterised by transcriptomic and cellular abundance differences in blood

**DOI:** 10.1101/2021.05.12.21257086

**Authors:** Jelmer Legebeke, Jenny Lord, Rebekah Penrice-Randal, Andres F. Vallejo, Stephen Poole, Nathan J. Brendish, Xiaofeng Dong, Catherine Hartley, John W. Holloway, Jane S. Lucas, Anthony P. Williams, Gabrielle Wheway, Fabio Strazzeri, Aaron Gardner, James P.R. Schofield, Paul J. Skipp, Julian A. Hiscox, Marta E. Polak, Tristan W. Clark, Diana Baralle

## Abstract

**Background:** The worldwide pandemic caused by SARS-CoV-2 has claimed millions of lives and has had a profound effect on global life. Understanding the pathogenicity of the virus and the body’s response to infection is crucial in improving patient management, prognosis, and therapeutic strategies. To address this, we performed functional transcriptomic profiling to better understand the generic and specific effects of SARS-CoV-2 infection.

**Methods:** Whole blood RNA sequencing was used to profile a well characterised cohort of patients hospitalised with COVID-19, during the first wave of the pandemic prior to the availability of approved COVID-19 treatments and who went on to survive or die of COVID-19, and patients hospitalised with influenza virus infection between 2017 and 2019. Clinical parameters between patient groups were compared, and several bioinformatic tools were used to assess differences in transcript abundances and cellular composition.

**Results:** The analyses revealed contrasting innate and adaptive immune programmes, with transcripts and cell subsets associated with the innate immune response elevated in patients with influenza, and those involved in the adaptive immune response elevated in patients with COVID-19. Topological analysis identified additional gene signatures that differentiated patients with COVID-19 from patients with influenza, including insulin resistance, mitochondrial oxidative stress and interferon signalling. An efficient adaptive immune response was furthermore associated with patient survival, while an inflammatory response predicted death in patients with COVID-19. A potential prognostic signature was found based on a selection of transcript abundances, associated with circulating immunoglobulins, nucleosome assembly, cytokine production and T cell activation, in the blood transcriptome of COVID-19 patients, upon admission to hospital, which can be used to stratify patients likely to survive or die.

**Conclusions:** The results identified distinct immunological signatures between SARS-CoV-2 and influenza, prognostic of disease progression and indicative of different targeted therapies. The altered transcript abundances associated with COVID-19 survivors can be used to predict more severe outcomes in patients with COVID-19.

## Introduction

The global pandemic caused by novel coronavirus SARS-CoV-2 emerged at the end of 2019 (1). By May 1^st^, 2021, more than 150 million people had been infected, leading to over 3 million deaths worldwide (2).

Previous studies investigating the differences between patients with COVID-19 or influenza on admission to hospital have found that both patient groups present with similar levels of systemic inflammation markers like C-reactive protein (CRP), white blood cell count (WBC), neutrophil count and neutrophil/lymphocyte (N/L) ratio (3). After admission patients hospitalised with COVID-19 were found to have a higher risk of developing respiratory distress, pulmonary embolism, septic shock and haemorrhagic strokes compared to influenza patients (4). In addition, the median length of stay in the intensive care unit was twice as high for COVID-19 patients compared to influenza patients, and COVID-19 patients were more likely to require mechanical ventilation (4). Furthermore, the in-hospital mortality for COVID-19 patients was 16.9% compared to 5.8% for influenza patients indicating a roughly three times higher relative risk of death for COVID-19 (4).

The viral immune response against influenza is well characterised (5). Briefly, initial defence involves cells of the innate immune system (e.g. macrophages, granulocytes and dendritic cells (DCs)), which release proinflammatory cytokines and type I interferons (IFN) to inhibit viral replication, recruit other immune cells to the site of infection, and stimulate the adaptive immune response. The adaptive immune response consists of a humoral and a cellular mediated immunity, initiated principally by virus-specific antibodies and T cells. Current understanding of the immune response specific to SARS-CoV-2 indicates that COVID-19 severity and duration are largely due to a total or early evasion of an innate immune and type I and type III interferon (IFN) responses (6–9), while patients infected with influenza are able to express type I and type II IFNs at a significantly higher concentration (3) which correlates with quicker recovery and decreased disease severity and mortality (10, 11). Consistent with this observation, early administration of inhaled recombinant IFN-beta for COVID-19 patients was associated with a lowered in-hospital mortality and quicker recovery (12, 13). Despite the reduced IFN response in patients with COVID-19 the expression of pro-inflammatory cytokines occurs for a prolonged time at similar levels with influenza patients (3), and interleukin (IL) −6 and IL-10 (14–16) and CCL3 (3) were associated with increased disease severity for COVID-19. The presence of CD4+ and CD8+ T cells, and antibodies were correlated with a positive patient outcome in the case of COVID-19 (17). This puts elderly patients at a higher risk due a smaller naïve T cell pool (18–20) and an absence of a pre-existing adaptive immunity (21) resulting in a potential delayed T cell response to a novel virus like SARS-CoV-2 (22). Delaying an adaptive immune response which, when combined with a high viral load, could lead to a poor outcome (23). As discussed by Sette and Crotty (24) an absent T cell response may cause an increased innate response attempting to control the virus resulting in an excessive lung immunopathology.

To investigate unique molecular features associated with COVID-19, a cohort of patients was identified from hospitalised individuals that were positive for SARS-CoV-2. As a comparator an equivalent group of patients hospitalised with influenza virus were identified. An extensive record of clinical parameters and peripheral blood, used for RNA-seq to obtain a global blood transcriptome overview, were taken at point of care and could therefore be correlated with any molecular signatures of disease. Through these side-by-side comparisons, we aim to identify distinct patterns of blood transcript abundances and cellular composition related to specific antiviral immune responses. Furthermore, we aim to identify a promising prognostic signature indicative of COVID-19 outcome.

## Materials & Methods

### Ethics and consent

The study was approved by the South Central - Hampshire A Research Ethics Committee (REC): REC reference 20/SC/0138 (March 16^th^, 2020) for the COVID-19 point of care (CoV-19POC) trial; and REC reference 17/SC/0368 (September 7^th^, 2017) for the FluPOC trial. For full inclusion and exclusion criteria details see (25) and (26). Patients gave written informed consent or consultee assent was obtained where patients were unable to give consent. Demographic and clinical data were collected at enrolment and outcome data from case note and electronic systems. ALEA and BC data management platforms were used for data capture and management.

### Study design and participants

All participants were recruited within the first 24 hours of admission to two large studies of molecular point-of-care testing (mPOCT) for respiratory viruses (CoV-19POC and FluPOC). Blood samples including whole blood in PAXgene Blood RNA tubes (BRT) (Preanalytix) were collected from SARS-CoV-2 positive patients and influenza positive patients, within 24 hours of enrolment, and stored at −80°C. The studies were prospectively registered with the ISRCTN trial registry: ISRCTN14966673 (COV-19POC) (March 18^th^, 2020), and ISRCTN17197293 (FluPOC) (November 13^th^, 2017). The COV-19POC study was a non-randomised interventional trial evaluating the clinical impact of mPOCT for SARS-CoV-2 in adult patients presenting to hospital with suspected COVID-19, using the QIAGEN QIAstat-Dx PCR testing platform with the QIAstat-Dx Respiratory SARS-CoV-2 Panel (27). The trial took place during the first wave of the pandemic, from 20th March to 29th April 2020, and prior to the availability of approved COVID-19 treatments. All patients were recruited from the Acute Medical Unit (AMU), Emergency Department (ED) or other acute areas of Southampton General Hospital. The FluPOC study was a multicentre randomised controlled trial evaluating the clinical impact of mPOCT for influenza in hospitalised adult patients with acute respiratory illness, during influenza season, using the BioFire FilmArray platform with the Respiratory Panel 2.1 (28). The trial took place during influenza seasons over the two winters of 2017/18 and 2018/19. All patients were recruited from the AMU and ED of Southampton General Hospital and Royal Hampshire County Hospital.

### Extraction of RNA from clinical samples and Illumina sequencing

Total RNA was extracted from PAXgene BRT using the PAXgene Blood RNA Kit (PreAnalytix), according to the manufacturer’s protocol at Containment Level 3 in a Tripass Class I hood. Extracted RNA was stored at −80°C until further use. Following the manufacturer’s protocols, total RNA was used as input material into the QIAseq FastSelect–rRNA/Globin Kit (Qiagen) protocol to remove cytoplasmic and mitochondrial rRNA and globin mRNA with a fragmentation time of 7 or 15 minutes. Subsequently the NEBNext® Ultra™ II Directional RNA Library Prep Kit for Illumina® (New England Biolabs) was used to generate the RNA libraries, followed by 11 or 13 cycles of amplification and purification using AMPure XP beads. Each library was quantified using Qubit and the size distribution assessed using the Agilent 2100 Bioanalyser and the final libraries were pooled in equimolar ratios. Libraries were sequenced using 150 bp paired-end reads on by an Illumina® NovaSeq 6000 (Illumina®, San Diego, USA). Raw fastq files were trimmed to remove Illumina adapter sequences using Cutadapt v1.2.1 (29). The option “−O 3” was set, so that the 3’ end of any reads which matched the adapter sequence with greater than 3 bp was trimmed off. The reads were further trimmed to remove low quality bases, using Sickle v1.200 (30) with a minimum window quality score of 20. After trimming, reads shorter than 10 bp were removed.

### Data QC and alignment

Quality control (QC) of read data was performed using FastQC (31) (v0.11.9) and compiled and visualised with MultiQC (32) (v1.5). Samples with <20 million total reads were excluded from further analysis. The STAR index was created with STAR’s (33) (v2.7.6a) genomeGenerate function using GRCh38.primary_assembly.genome.fa and gencode.v34.annotation.gtf (34) (both downloaded from GENCODE), with –sjdbOverhang 149 and all other settings as default. Individual fastq files were aligned using the --twopassMode Basic flag, with the following parameters specified (following ENCODE standard options): --outSAMmapqUnique 60, outFilterType BySJout, -- outFilterMultimapNmax 20, --alignSJoverhangMin 8, --outFilterMismatchNmax 999, -- outFilterMismatchNoverReadLmax 0.04, --alignIntronMin 20, --alignIntronMax 1000000, -- alignMatesGapMax 1000000 and all other options as default. For rMATs (35) (v4.1.0) analysis, STAR was run again as before, but with the addition of --alignEndsType EndToEnd. Samtools (36) (v1.8) was used to sort and index the aligned data.

### Comparisons of baseline clinical characteristics

Baseline clinical characteristics of the patient groups were assessed using R (37) (v4.0.2) and RStudio (38) (v1.3.959) for comparisons between COVID-19 versus influenza, and COVID-19 survivors versus non-survivors. Extreme outliers (values < Q1 - 3 interquartile range, or > Q3 + 3 interquartile range) were identified with the R package rstatix (39) (v0.7.0) and removed. Statistical testing was performed including a Shapiro-Wilk test to assess for data normality followed with either an unpaired parametric T-test (Shapiro-Wilk test p-value > 0.05) or an unpaired non-parametric Wilcoxon test (Shapiro-Wilk test p-value < 0.05) for continuous data, or a Chi-square test for categorical data. The R package Table1 (40) (v1.3) was used to plot the baseline clinical characteristics.

### Systems immunology-based analysis of blood transcript modules

Blood transcript module (BTM) analysis was performed with molecular signatures derived from 5 vaccine trials (41) as a reference dataset, and BTM activity was calculated using the BTM package (41) (v1.015) in Python (42) (v3.7.2) using the normalized counts as input. Module enrichment significance was calculated using CAMERA (43) (v3.46.0). The significance threshold for the linear model was set at false discovery rate (FDR) 0.05 for the comparison between patients with COVID-19 or influenza.

### Unbiased gene clustering analysis

Gene co-expression analysis was performed with BioLayout (44) (v3.4) using a correlation value of 0.95, other settings were kept at default. Clusters were manually assessed to determine gene expression differences depending on for example patient cohort. Genes were subsequently analysed with ToppGene (45) gene list enrichment analysis using the default settings.

### Differential gene expression analysis between patient groups

HTSeq (46) (v0.11.2) count was used to assign counts to RNA-seq reads in the Samtools sorted BAM file using GENCODE v34 annotation. Parameters used for HTSeq were --format=bam, --order=pos, -- stranded=reverse, --type=exon and the other options were kept at default. EdgeR (47) (v3.30.3) was used for differential gene expression analysis with R (v4.0.2) in RStudio (v1.3.959). Genes with low counts across all libraries were filtered out using the filterByExpr command. Filtered gene counts were normalised using the trimmed mean of M-values (TMM) method. Differentially expressed genes were identified, after fitting the negative binomial models and obtaining dispersion estimates, using the exact test and using a threshold criteria of FDR p-value < 0.05 and log2 fold change < −1 and > 1. Genes which were within the threshold criteria were used for ToppGene gene list enrichment analysis. A principal component analysis (PCA) graph was constructed based on all differentially expressed genes to assess sample clustering.

### Assessment of difference in adaptive immune response related gene expression

A higher abundance of transcripts from 83 immunoglobulin genes, overlapping with the genes in the Gene Ontology (GO) (48, 49) biological process term ‘adaptive immune response’ (**Additional file 1**), was found in patients with COVID-19 compared to influenza. To assess gene transcript abundance differences for these 83 genes in each patient a heatmap was generated and Z-scores were summed to give an overall positive (high) or negative (low) total Z-score. Patient baseline clinical characteristics were explored, as above, for any explanatory factors for the involvement of a high or low total Z-score between patients with COVID-19 or influenza, and those that survived COVID-19 versus those that died within 30 days of hospital admission. Metadata comparison plots were made with the R package ggplot2 (50) (3.3.2) and statistical testing with the R package ggpubr (51) (v0.4.0).

### Topological mapping of global gene patterns

TopMD Pathway Analysis (52) was conducted using the differential transcript abundances identified by differential gene expression analysis, generating a map of the differentially activated pathways between all patients with COVID-19 or influenza. The TopMD pathway algorithm measures the geometrical and topological properties of global differential gene expression embedded on a gene interaction network (53). This enables plotting and measurement of the differentially activated pathways through extrapolation of groups of mechanistically related genes, called TopMD pathways. TopMD pathways possess a natural hierarchical structure and can be analysed for enriched GO terms, by chi-square test.

### Assessment of differential splicing between patient groups

Three different tools were used to assess differential gene splicing between patients with COVID-19 or influenza, and COVID-19 survivors or non-survivors after 30 days of hospital admission. rMATs (35) (v4.1.0) was run using BAM files with soft clipping suppressed, generated with STAR and GENCODE v34 gene annotation. Additional settings used were -t paired, --readLength 150 and --libType fr-firststrand. Results were filtered for FDR p-value < 0.05. LeafCutter (54) (v0.2.9) was run in stages following the Differential Splicing protocol (55) (bam2junc.sh generated junction files from BAMs, leafcutter_cluster.py grouped junctions into clusters, leafcutter_ds.R tested for differential splicing, all with default settings, except –min_samples_per_intron was set to be approximately 60% of the smaller group size for each comparison (46 for COVID-19 vs influenza, 9 for COVID-19 survivors vs non-survivors), and results were filtered to exclude events with delta PSI <10%, based on recommendations (56). The LeafViz script (57), prepare_results.R was used to generate a data table from which gene names for significant events were extracted, while the map_clusters_to_genes R function was used to assign genes to non-significant tested events. Overlap between LeafCutter differentially spliced and EdgeR differentially expressed genes was tested for significance using Fisher’s Exact Test (fisher.test in R (v3.5.1) using a 2×2 contingency table and two.sided alternative hypothesis). MAJIQ (58) (v2.2) was run in two stages (majiq build and majiq deltapsi) with default settings, and results were filtered (delta PSI >20%, probability >0.95) using Voila (58) (v2.0).

### *In silico* immune profiling predicting immune cell levels between patient groups

Relative abundance of 22 immune cell types and their statistical significance was deconvoluted from whole blood using the reference gene signature matrix (LM22) using CIBERSORTx (59). CIBERSORTx analysis was conducted on the CIBERSORTx website (60) using 100 permutations. Immune cell distribution between the groups were compared by Mann–Whitney test.

### Identification of immune signatures as a predictor for COVID-19 outcome

Transcript to transcript gene co-expression network analysis with BioLayout 3D (v3.4) (Pearson coefficient 0.85, MCL=1.7) assembled 537 genes differentially expressed (EdgeR, FDR < 0.5 and |log2 fold change > 1|) in blood taken on admission between patients with COVID-19 who either survived or died of COVID-19 within 30 days of admission to hospital. Combinations of 100 genes from the top 4 clusters were assessed as predictor variables for outcome using Boosted Logistic Regression, Bayesian Generalised Linear and RandomForest models within SIMON (61) (v0.2.1) installed with Docker (62) (v20.10.2). TMM normalised gene expression data was centred and scaled. Covariant features were removed based on correlation analysis. Samples were randomly split into train:test subsets at the ratio 75%:25%.

## Results

### Number of participants

In total RNA-seq was done for 80 patients with COVID-19 and 88 patients with influenza. Five patients with influenza failed QC (read count < 20M) leaving 83 patients with influenza for analysis, of which 76% were infected with the influenza A virus and 22% with influenza B virus. Two patients with COVID-19 were identified by PCA as outliers, subsequent assessment revealed an elevated white blood cell and lymphocyte count caused by pre-existing underlying chronic lymphocytic leukaemia, and these patients were excluded from further analyses (**Supplementary figure 1**). This left 78 patients with COVID-19, of whom 62 survived and 16 died within 30 days of hospital admission, and 83 patients with influenza.

### Clinical differences

The baseline clinical characteristics of the patients used in this study for the comparison between influenza and COVID-19 were assessed and no differences in distribution of sex or age were detected between patient groups, however, more patients with influenza were of White British ethnicity (p-value 1.12×10^−05^) and more were current smokers (p-value 9.07×10^−05^). There were also differences in the proportion of cases with underlying comorbidities, with patients with COVID-19 more commonly having hypertension (p-value 1.42×10^−02^), liver disease (p-value 3.63×10^−02^) and diabetes mellitus (p-value 6.44×10^−03^) than those with influenza. However, underlying respiratory disease was more common in patients with influenza (p-value 1.22×10^−03^). Patients with COVID-19 generally exhibited more severe clinical symptoms. While the National Early Warning Score 2 (NEWS2) was not different between patients with COVID-19 or influenza, patients with COVID-19 had a higher respiratory rate (p-value 2.79×10^−02^) and a greater proportion of patients with COVID-19 were on supplementary oxygen at hospital admission (p-value 6.81×10^−03^). Laboratory results indicated higher levels of C-reactive protein (p-value 1.73×10^−03^) and lymphocytes (p-value 2.76×10^−02^) in patients with COVID-19. Furthermore, COVID-19 patients had a longer duration of symptoms prior to presentation to hospital (p-value 1.17×10^−05^) and once admitted a longer length of stay (p-value 5.51×10^−10^). Longer stay time was associated with increased 30 day mortality after hospital admission and patients with COVID-19 were more likely to have died compared to patients with influenza (p-value 4.42×10^−05^) (**Table 1**).

**Table 1:** Baseline clinical characteristics and outcomes of hospitalised patients with COVID-19 or influenza.

| Baseline demographic data |  |  |  | Comorbidity (cont.) |  |  |  |
| --- | --- | --- | --- | --- | --- | --- | --- |
|  | COVID-19<br>(N=78) | Influenza<br>(N=83) | P-value |  | COVID-19<br>(N=78) | Influenza<br>(N=83) | P-value |
| **Sex** |  |  |  | **Other respiratory disease** |  |  |  |
| Female | 26 (33.3%) | 36 (43.4%) | 0.252 | Yes | 21 (26.9%) | 44 (53.0%) | 0.00122 |
| Male | 52 (66.7%) | 47 (56.6%) |  | Unknown | 3 (3.8%) | 0 (0%) |  |
| **Age (years)** |  |  |  | **Clinical observations** |  |  |  |
| Mean (SD) | 60.9 (18.0) | 57.8 (18.4) | 0.367 |  | COVID-19<br>(N=78) | Influenza<br>(N=83) | P-value |
| **Ethnic category (Code 2001)** |  |  |  | **Heart rate (BPM)** |  |  |  |
| White - British | 47 (60.3%) | 79 (95.2%) | 1.12e-05 | Mean (SD) | 97.3 (17.1) | 101 (23.0) | 0.39 |
| Asian - Indian | 3 (3.8%) | 0 (0%) |  | **Systolic blood pressure (mmHg)** |  |  |  |
| Black - African | 6 (7.7%) | 0 (0%) |  | Mean (SD) | 133 (19.9) | 132 (23.6) | 0.993 |
| Black - Caribbean | 2 (2.6%) | 0 (0%) |  | **Respiratory rate (bpm)** |  |  |  |
| Other White background | 6 (7.7%) | 3 (3.6%) |  | Mean (SD) | 26.6 (7.73) | 23.8 (5.96) | 0.0279 |
| Other Asian background | 13 (16.7%) | 0 (0%) |  | **Temperature (Celsius)** |  |  |  |
| Mixed | 0 (0%) | 1 (1.2%) |  | Mean (SD) | 37.4 (1.01) | 37.7 (1.13) | 0.0822 |
| Not stated | 1 (1.3%) | 0 (0%) |  | **Oxygen saturation (%)** |  |  |  |
| **Current smoking status** |  |  |  | Mean (SD) | 94.3 (3.75) | 94.8 (3.41) | 0.548 |
| Yes | 4 (5.1%) | 21 (25.3%) | 9.07e-05 | **Supplementary O2, n (%)** |  |  |  |
| No | 67 (85.9%) | 62 (74.7%) |  | Yes | 37 (47.4%) | 21 (25.3%) | 0.00681 |
| Unknown | 7 (9.0%) | 0 (0%) |  | No | 41 (52.6%) | 61 (73.5%) |  |
| **Symptom duration (days)** |  |  |  | **NEWS2** |  |  |  |
| Median [Min, Max] | 7.00 [0, 21.0] | 4.00 [1.00, 10.0] | 1.17e-05 | Mean (SD) | 5.28 (2.78) | 4.79 (2.57) | 0.171 |
| **Comorbidity** |  |  |  | **Laboratory results** |  |  |  |
|  | COVID-19<br>(N=78) | Influenza<br>(N=83) | P-value |  | COVID-19<br>(N=78) | Influenza<br>(N=83) | P-value |
| **Hypertension** |  |  |  | **White blood cell count** |  |  |  |
| Yes | 29 (37.2%) | 20 (24.1%) | 0.0142 | Mean (SD) | 8.73 (4.29) | 8.64 (3.89) | 0.913 |
| Unknown | 4 (5.1%) | 0 (0%) |  | **Neutrophil cell count** |  |  |  |
| **Cardiovascular disease** |  |  |  | Mean (SD) | 7.06 (4.07) | 6.93 (3.67) | 0.895 |
| Yes | 16 (20.5%) | 14 (16.9%) | 0.152 | **Lymphocyte cell count** |  |  |  |
| Unknown | 3 (3.8%) | 0 (0%) |  | Mean (SD) | 1.01 (0.411) | 0.908 (0.541) | 0.0276 |
| **Renal disease** |  |  |  | **C-reactive protein** |  |  |  |
| Yes | 6 (7.7%) | 4 (4.8%) | 0.141 | Mean (SD) | 131 (110) | 80.2 (78.9) | 0.00173 |
| Unknown | 3 (3.8%) | 0 (0%) |  | **Outcomes** |  |  |  |
| **Liver disease** |  |  |  |  | COVID-19<br>(N=78) | Influenza<br>(N=83) | P-value |
| Yes | 3 (3.8%) | 0 (0%) | 0.0363 | **Length of stay (days)** |  |  |  |
| Unknown | 3 (3.8%) | 0 (0%) |  | Mean (SD) | 10.5 (9.51) | 3.39 (2.92) | 5.51e-10 |
| **Diabetes mellitus** |  |  |  | **Died within 30 days after admission** |  |  |  |
| Yes | 19 (24.4%) | 8 (9.6%) | 0.00644 | Yes | 16 (20.5%) | 0 (0%) | 4.42e-05 |
| Unknown | 3 (3.8%) | 0 (0%) |  | No | 62 (79.5%) | 83 (100%) |  |
| **Active malignancy** |  |  |  |  |  |  |  |
| Yes | 6 (7.7%) | 6 (7.2%) | 0.193 |  |  |  |  |
| Unknown | 3 (3.8%) | 0 (0%) |  |  |  |  |  |
| **Immunosuppressed** |  |  |  |  |  |  |  |
| Yes | 4 (5.1%) | 5 (6.0%) | 0.111 |  |  |  |  |
| Unknown | 4 (5.1%) | 0 (0%) |  |  |  |  |  |

Comparisons are given between patients with COVID-19 or influenza for baseline demographic data, patient outcome, clinical observations, laboratory results and known patient comorbidity. Laboratory results were done on peripheral blood taken on admission to hospital. Similarly, clinical observations were recorded on hospital admission. Statistical testing was done with a Shapiro-Wilk test for data normality followed with either an unpaired parametric T-test or an unpaired non-parametric Wilcoxon test for continuous data, or a Chi-square test for categorical data.

Between patients with COVID-19 who survived and those who died, a fatal outcome occurred in older patients (p-value 2.58×10^−09^). COVID-19 non-survivors also had a shorter duration of symptoms before being admitted to hospital (p-value 5.38×10^−03^). COVID-19 non-survivors more commonly had underlying comorbidities including hypertension (p-value 1.93×10^−03^), cardiovascular disease (p-value 3.97×10^03^), diabetes mellitus (p-value 2.31−10^−02^) and underlying respiratory disease (p-value 1.06×10^−02^). While the NEWS2 scores were not different, COVID-19 survivors had a higher heart rates than COVID-19 non-survivors (p-value 9.27×10^−03^). Laboratory results showed an increase of white blood cell count (p-value 3.83×10^−02^), total protein levels (p-value 2.5×10^−03^), creatinine (p-value 3.87×10^−02^), alanine aminotransferase levels (p-value 2.85×10^−02^), troponin levels (p-value 2.37×10^−04^), tumour necrosis factor α (TNFα) (p-value 1.43×10^−02^), interleukin (IL)-6 levels (p-value 2.78−10^−03^), IL-8 (p-value 2.24×10^−02^), IL-1β (p-value 3.78×10^−02^) and IL-10 (p-value 7.51×10^−02^) in COVID-19 non-survivors. Patient outcome and length of hospital stay were different due to separation based on patient survival (**Table 2**).

**Table 2:** Baseline clinical characteristics and outcomes of hospitalised COVID-19 patients: survivors versus non-survivors.

| Baseline demographic data |  |  |  | Laboratory results |  |  |  |
| --- | --- | --- | --- | --- | --- | --- | --- |
|  | Non-survivor<br>(N=16) | Survivor<br>(N=62) | P-value |  | Non-survivor<br>(N=16) | Survivor<br>(N=62) | P-value |
| Sex, n (%) |  |  |  | Haemoglobin count (g/L) |  |  |  |
| Female | 7 (43.8%) | 19 (30.6%) | 0.488 | Mean (SD) | 128 (21.3) | 138 (20.7) | 0.144 |
| Male | 9 (56.2%) | 43 (69.4%) |  | White blood cell count (10<sup>9</sup>/L) |  |  |  |
| Age (years) |  |  |  | Mean (SD) | 10.4 (4.27) | 8.31 (4.23) | 0.0383 |
| Mean (SD) | 81.6 (10.4) | 55.6 (15.6) | 2.58e-09 | Platelet count (10<sup>9</sup>/L) |  |  |  |
| Ethnic category (Code 2001), n (%) |  |  |  | Mean (SD) | 231 (83.9) | 249 (90.0) | 0.38 |
| White - British | 14 (87.5%) | 33 (53.2%) | 0.203 | Neutrophil cell count (10<sup>9</sup>/L) |  |  |  |
| Asian - Indian | 1 (6.2%) | 2 (3.2%) |  | Mean (SD) | 8.73 (4.15) | 6.66 (3.98) | 0.063 |
| Black - African | 1 (6.2%) | 5 (8.1%) |  | Lymphocyte cell count (10<sup>9</sup>/L) |  |  |  |
| Black - Caribbean | 0 (0%) | 2 (3.2%) |  | Mean (SD) | 0.900 (0.419) | 1.04 (0.409) | 0.142 |
| Other White background | 0 (0%) | 6 (9.7%) |  | Sodium level (mmol/L) |  |  |  |
| Other Asian background | 0 (0%) | 13 (21.0%) |  | Mean (SD) | 133 (7.01) | 136 (3.90) | 0.0878 |
| Not stated | 0 (0%) | 1 (1.6%) |  | Potassium level (mmol/L) |  |  |  |
| Current smoking status, n (%) |  |  |  | Mean (SD) | 4.15 (0.971) | 4.02 (0.473) | 0.824 |
| Yes | 0 (0%) | 4 (6.5%) | 0.0291 | Urea levels (mmol/L) |  |  |  |
| No | 12 (75.0%) | 55 (88.7%) |  | Mean (SD) | 11.6 (5.98) | 6.61 (3.32) | 0.0025 |
| Unknown | 4 (25.0%) | 3 (4.8%) |  | Creatinine level (μmol/L) |  |  |  |
| Symptom duration (days) |  |  |  | Mean (SD) | 128 (66.7) | 83.4 (25.2) | 0.0387 |
| Median [Min, Max] | 2.00 [0, 14.0] | 7.00 [0, 21.0] | 0.00538 | Albumin level (g/L) |  |  |  |
| Comorbidity |  |  |  | Mean (SD) | 33.9 (4.66) | 32.8 (4.78) | 0.443 |
|  | Non-survivor<br>(N=16) | Survivor<br>(N=62) | P-value | Bilirubin level (μmol/L) |  |  |  |
| Hypertension, n (%) |  |  |  | Mean (SD) | 12.0 (6.06) | 11.1 (4.26) | 0.965 |
| Yes | 12 (75.0%) | 17 (27.4%) | 0.00193 | Alanine aminotransferase level (units/L) |  |  |  |
| Unknown | 0 (0%) | 4 (6.5%) |  | Mean (SD) | 37.0 (37.3) | 54.1 (43.4) | 0.0285 |
| Cardiovascular disease, n (%) |  |  |  | Alkaline phosphatase level (units/L) |  |  |  |
| Yes | 8 (50.0%) | 8 (12.9%) | 0.00397 | Mean (SD) | 93.2 (46.0) | 95.2 (48.1) | 0.922 |
| Unknown | 0 (0%) | 3 (4.8%) |  | Total protein level (g/L) |  |  |  |
| Renal disease, n (%) |  |  |  | Mean (SD) | 72.7 (9.98) | 69.9 (6.26) | 0.367 |
| Yes | 3 (18.8%) | 3 (4.8%) | 0.129 | Lactate dehydrogenase level (units/L) |  |  |  |
| Unknown | 0 (0%) | 3 (4.8%) |  | Mean (SD) | 841 (357) | 914 (486) | 0.864 |
| Liver disease, n (%) |  |  |  | Ferritin level (mmol/L) |  |  |  |
| Yes | 0 (0%) | 3 (4.8%) | 0.432 | Mean (SD) | 1420 (2020) | 974 (794) | 0.841 |
| Unknown | 0 (0%) | 3 (4.8%) |  | Troponin level (ng/L) |  |  |  |
| Diabetes mellitus, n (%) |  |  |  | Mean (SD) | 164 (194) | 9.55 (6.67) | 0.000237 |
| Yes | 8 (50.0%) | 11 (17.7%) | 0.0231 | C-reactive protein (mg/L) |  |  |  |
| Unknown | 0 (0%) | 3 (4.8%) |  | Mean (SD) | 172 (165) | 121 (90.7) | 0.662 |
| Active malignancy, n (%) |  |  |  | IL-1B level (pg/ml) |  |  |  |
| Yes | 3 (18.8%) | 3 (4.8%) | 0.129 | Mean (SD) | 0.620 (0.474) | 0.378 (0.200) | 0.0378 |
| Unknown | 0 (0%) | 3 (4.8%) |  | IL-6 level (pg/ml) |  |  |  |
| Immunosuppressed, n (%) |  |  |  | Mean (SD) | 174 (142) | 59.9 (47.8) | 0.00278 |
| Yes | 1 (6.2%) | 3 (4.8%) | 0.946 | IL-8 level (pg/ml) |  |  |  |
| Unknown | 1 (6.2%) | 3 (4.8%) |  | Mean (SD) | 58.6 (29.0) | 41.2 (26.5) | 0.0224 |
| Other respiratory disease, n (%) |  |  |  | IL-10 level (pg/ml) |  |  |  |
| Yes | 9 (56.2%) | 12 (19.4%) | 0.0106 | Mean (SD) | 39.5 (36.7) | 15.7 (9.35) | 0.00181 |
| Unknown | 0 (0%) | 3 (4.8%) |  | IL-33 level (pg/ml) |  |  |  |
| Clinical observations |  |  |  | Mean (SD) | 0.543 (0.387) | 0.340 (0.277) | 0.0751 |
|  | Non-survivor<br>(N=16) | Survivor<br>(N=62) | P-value | TNFα level (pg/ml) |  |  |  |
| Heart rate (beats-per-minute) |  |  |  | Mean (SD) | 30.1 (15.6) | 19.3 (6.87) | 0.0143 |
| Mean (SD) | 87.6 (15.1) | 99.9 (16.8) | 0.00927 | GM-CSF level (pg/ml) |  |  |  |
| Systolic blood pressure (mmHg) |  |  |  | Mean (SD) | 2.08 (2.61) | 1.48 (0.972) | 0.753 |
| Mean (SD) | 132 (29.8) | 133 (16.8) | 0.853 | IFNγ level (pg/ml) |  |  |  |
| Respiratory rate (breaths-per-minute) |  |  |  | Mean (SD) | 35.3 (71.7) | 26.6 (55.5) | 0.313 |
| Mean (SD) | 27.8 (7.57) | 26.3 (7.80) | 0.337 | Outcomes |  |  |  |
| Temperature (Celsius) |  |  |  |  | Non-survivor<br>(N=16) | Survivor<br>(N=62) | P-value |
| Mean (SD) | 37.3 (1.14) | 37.4 (0.978) | 0.804 | Length of stay (days) |  |  |  |
| Oxygen saturation (%) |  |  |  | Mean (SD) | 4.93 (2.34) | 11.9 (10.1) | 0.00529 |
| Mean (SD) | 93.4 (6.12) | 94.6 (2.83) | 0.643 | Median [Min, Max] | 5.00 [1.00, 8.00] | 9.00 [0, 44.0] |  |
| Supplementary O2, n (%) |  |  |  | Missing | 1 (6.2%) | 3 (4.8%) |  |
| Yes | 8 (50.0%) | 29 (46.8%) | 1 | Died within 30 days after admission, n (%) |  |  |  |
| No | 8 (50.0%) | 33 (53.2%) |  | Yes | 16 (100%) | 0 (0%) | <2e-16 |
| National Early Warning Score 2 |  |  |  | No | 0 (0%) | 62 (100%) |  |
| Mean (SD) | 5.40 (2.44) | 5.25 (2.88) | 0.906 |  |  |  |  |

Comparisons are given between COVID-19 survivors and non-survivors for baseline demographic data, patient outcome, clinical observations, laboratory results and known patient comorbidity. Laboratory results were done on peripheral blood taken on admission to hospital. Similarly, clinical observations were recorded on hospital admission. Statistical testing was done with a Shapiro-Wilk test for data normality followed with either an unpaired parametric T-test or an unpaired non-parametric Wilcoxon test for continuous data, or a Chi-square test for categorical data.

### Molecular differences

RNA-seq was used to investigate potential blood transcriptomic signatures of immune activation between patients infected with SARS-CoV-2 versus influenza, and COVID-19 survivors versus those that died. A median of 60.4 million reads in patients with COVID-19, and 58.9 million reads in patients with influenza was obtained (**Supplementary figure 2A**). In patients who died of COVID-19 a median of 55.7 million reads was obtained and for COVID-19 survivors the median was 62.6 million reads (**Supplementary figure 2B**). Clustering analysis between patients with COVID-19 or influenza indicated a homogeneity of blood transcriptome profiles suggesting any variation between groups to be subtle (**Supplementary figure 3A**). A partial separation was found between patients who survived or died of COVID-19 based on patient outcome after 30 days of hospital admission, indicative of a larger variation in the blood transcriptome (**Supplementary figure 3B**).

### Contrasting innate and adaptive immune programmes

Previous studies have suggested that severe COVID-19 is associated with aberrant immune pathology (63, 64), and therefore BTM analysis and gene co-expression analysis were used to investigate the balance between the innate and adaptive response in patients with either COVID-19 or influenza virus and to identify patterns of changes associated with each arm of the immune system. A systems immunology-based analysis of BTMs between patients with COVID-19 or influenza revealed several differences (**Figure 1**). For the upregulated BTMs in COVID-19, signatures were observed related to the cell cycle and adaptive immune response, primarily CD4+ T cells, B cells, plasma cells and immunoglobulins. In contrast, the downregulated BTMs showed signatures associated with monocytes, inflammatory signalling and an innate antiviral and type I IFN response.

**Figure 1:**
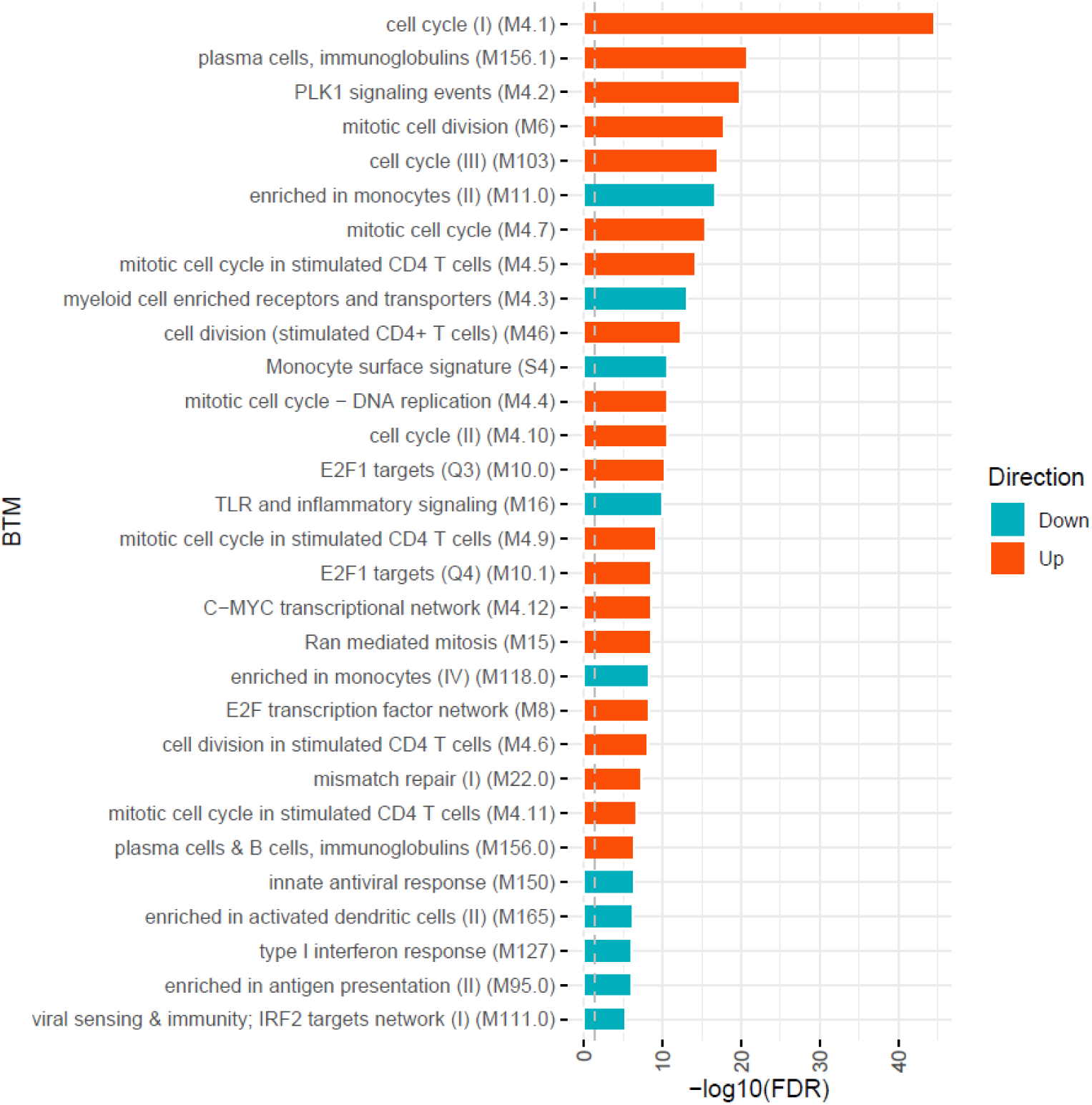
Blood transcript module (BTM) analysis between patients with COVID-19 or influenza. Upregulated signatures in COVID-19 patients are associated with cell cycle and an adaptive immune response, primarily CD4+ T cells, B cells, plasma cells and immunoglobulins. While the downregulated signatures, associated with influenza patients, are involved with monocytes, inflammatory signalling and an innate antiviral and type I interferon response.

Gene co-expression analysis was done on a total of 4,093 transcript abundances for unbiased gene clustering between patients with COVID-19 or influenza (TMM normalised counts per million EdgeR FDR p-value <0.05) and identified 50 clusters of 4 or more genes (BioLayout 3D, Pearson R >= 0.85, MCL = 1.7). These clusters are clearly separated into groups comprising increased transcript abundances in blood of patients with influenza or COVID-19 (**Figure 2**) and the top 12 clusters are shown in **Table 3**.

**Figure 2:**
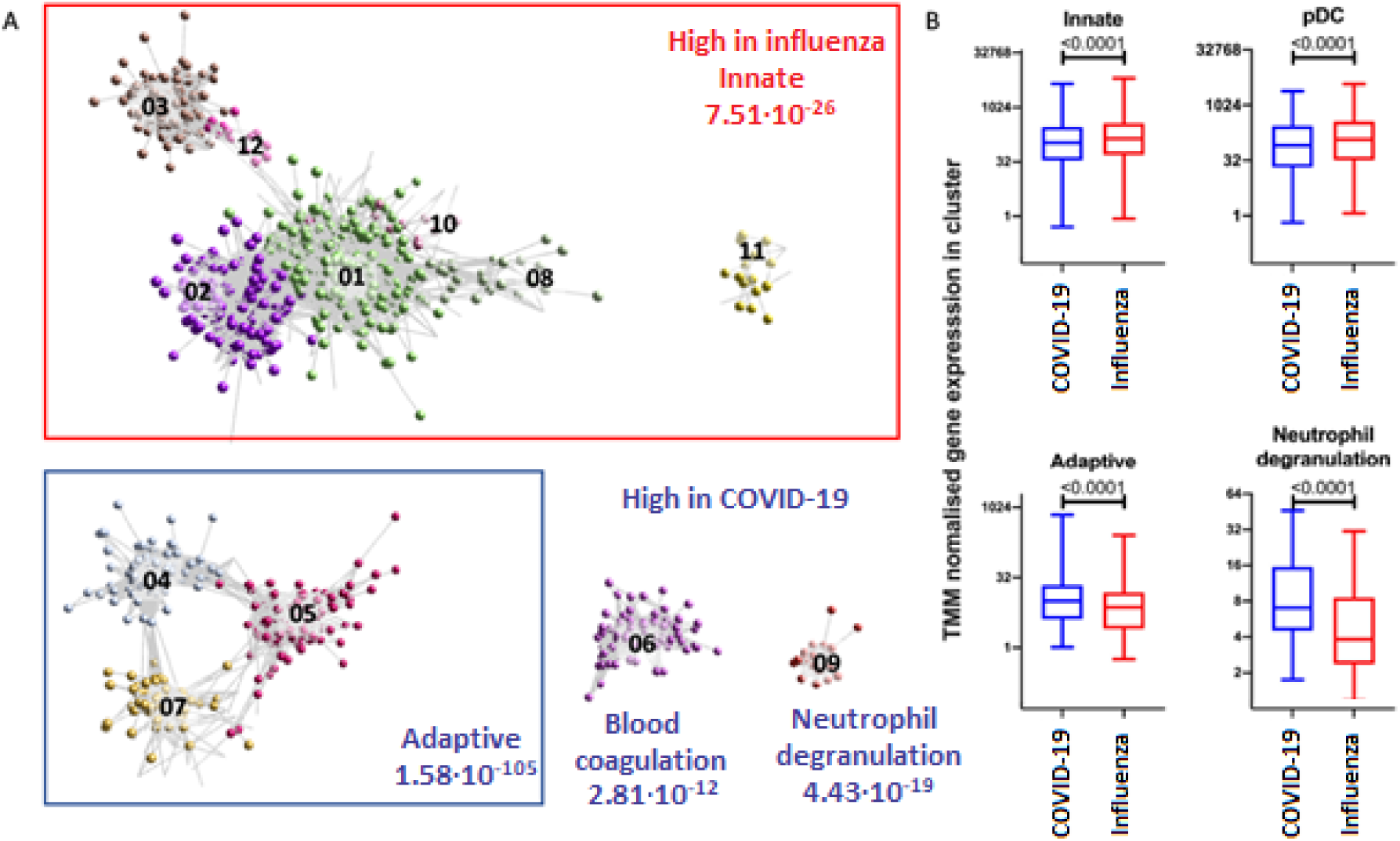
Top 12 clusters identified with BioLayout. A) Enrichment of gene clusters in blood of patients with influenza (annotated in red) and COVID-19 (annotated in blue). Increased abundances of gene transcripts in influenza patients are involved with an innate immune response, while in COVID-19 clusters are involved with an adaptive immune response, blood coagulation and neutrophil degranulation. B) After TMM normalisation a significant difference in gene clusters between patients with influenza or COVID-19 was detected. The abundance of gene transcripts involved with an innate immune response and plasmacytoid dendritic cell were observed to be higher in influenza patients. In contrast, the abundance of gene transcripts involved with an adaptive immune response and neutrophil degranulation was higher in COVID-19 patients.

**Table 3:** Summary of the top 12 BioLayout clusters.

| Cluster | No. of genes | Cell type (FDR p-value) | Top biological process (FDR p-value) | Disease |
| --- | --- | --- | --- | --- |
| 1 | 362 | Myeloid ($1.20 \times 10^{-24}$) | Cell activation ($5.16 \times 10^{-13}$) | Influenza |
| 2 | 264 | Plasmacytoid dendritic cell ($4.17 \times 10^{-22}$) | Defence response to virus ($1.34 \times 10^{-37}$) | Influenza |
| 3 | 166 | Erythroblast ($5.31 \times 10^{-20}$) | Erythrocyte differentiation ($1.70 \times 10^{-05}$) | Influenza |
| 4 | 140 | Progenitor B cell / T cell ($1.28 \times 10^{-131}$) | Mitotic cell cycle ($3.97 \times 10^{-57}$) | COVID-19 |
| 5 | 100 | Progenitor pluripotent cells ($1.38 \times 10^{-02}$) | Translation ($8.48 \times 10^{-04}$) | COVID-19 |
| 6 | 96 | Megakaryocytes / platelets ($3.30 \times 10^{-92}$) | Blood coagulation ($2.84 \times 10^{-12}$) | COVID-19 |
| 7 | 64 | Plasma cells ($1.27 \times 10^{-28}$) | Response to stress ($6.41 \times 10^{-09}$) | COVID-19 |
| 8 | 29 | Myeloid cells ($2.57 \times 10^{-03}$) | Myeloid leukocyte activation ($4.15 \times 10^{-04}$) | Influenza |
| 9 | 20 | Neutrophils ($1.11 \times 10^{-03}$) | Neutrophil degranulation ($4.43 \times 10^{-19}$) | COVID-19 |
| 10 | 18 | Antigen presenting cells ($2.21 \times 10^{-03}$) | Th1 stimulation ($4.53 \times 10^{-03}$) | Influenza |
| 11 | 16 | Dendritic cells ($4.32 \times 10^{-04}$) | Cell morphogenesis ($1.37 \times 10^{-02}$) | Influenza |
| 12 | 14 | Not specified | Histone modification ($3.55 \times 10^{-02}$) | Influenza |

Gene clusters were identified with BioLayout ( $r=0.85$ , $MCL = 1.7$ ). For each cluster the number of genes, predicted cell type and top biological process are given and whether that cluster was enriched in patients with COVID-19 or influenza.

Interestingly, an increased abundance of gene transcripts in patients with COVID-19 are involved in adaptive immunity, pointing to activation/priming of T cells and B cells, including induction of proliferation (cluster 4, FDR p-value 3.97×10^−57^), Additionally, an increased abundance of gene transcripts encoding neutrophil degranulation (cluster 9) and blood coagulation (cluster 6) clearly differentiated patients with COVID-19 from patients with influenza (FDR p-value 4.33×10^−19^ and FDR p-value 2.84×10^−12^ respectively). In contrast, an decreased abundance of gene transcripts in the blood transcriptome of patients with COVID-19 in comparison to patients with influenza were associated with innate immunity, including biological processes involved with defence response to virus (cluster 2) (FDR p-value 1.34×10^−37^), type 1 helper T cell stimulation (cluster 10) (FDR p-value 4.53×10^−03^), dendritic cell morphogenesis (cluster 11) (FDR p-value 1.37×10^−02^), and myeloid cell activation (clusters 1 and 8) (FDR p-value 5.16×10^−13^ and FDR p-value 4.15×10^−04^ respectively). Importantly, the largest decrease of transcript abundances in patients with COVID-19 comprised genes expressed in plasmacytoid dendritic cells (pDC) (FDR p-value 4.17×10^−22^), indicating impaired immune responses to viruses (FDR p-value 1.34×10^−37^) and impaired IFN signalling (FDR p-value 5.56×10^−30^). This was suggestive of contrasting innate and adaptive immune programmes between the different infections and these were further investigated.

### High abundance of immunoglobulin genes associated COVID-19

A total of 20,542 abundance measures of gene transcripts were obtained after filtering out transcripts with low counts, of which 4,094 transcripts were found to be significantly different between patients with COVID-19 or influenza (FDR p-value < 0.05) of which, 197 transcripts exceeded a log2 fold change of < −1 or >1, with 126 transcripts showing higher abundance in patients with COVID-19 and 71 transcripts showing higher abundance in patients with influenza (**Figure 3A** and **Additional file 2**). Complimentary to the findings from gene co-expression analysis, the transcripts with increased abundance in patients with COVID-19 were found to be involved with humoral immune response, complement activation and B cell mediated immunity (**Figure 3B**).

**Figure 3:**
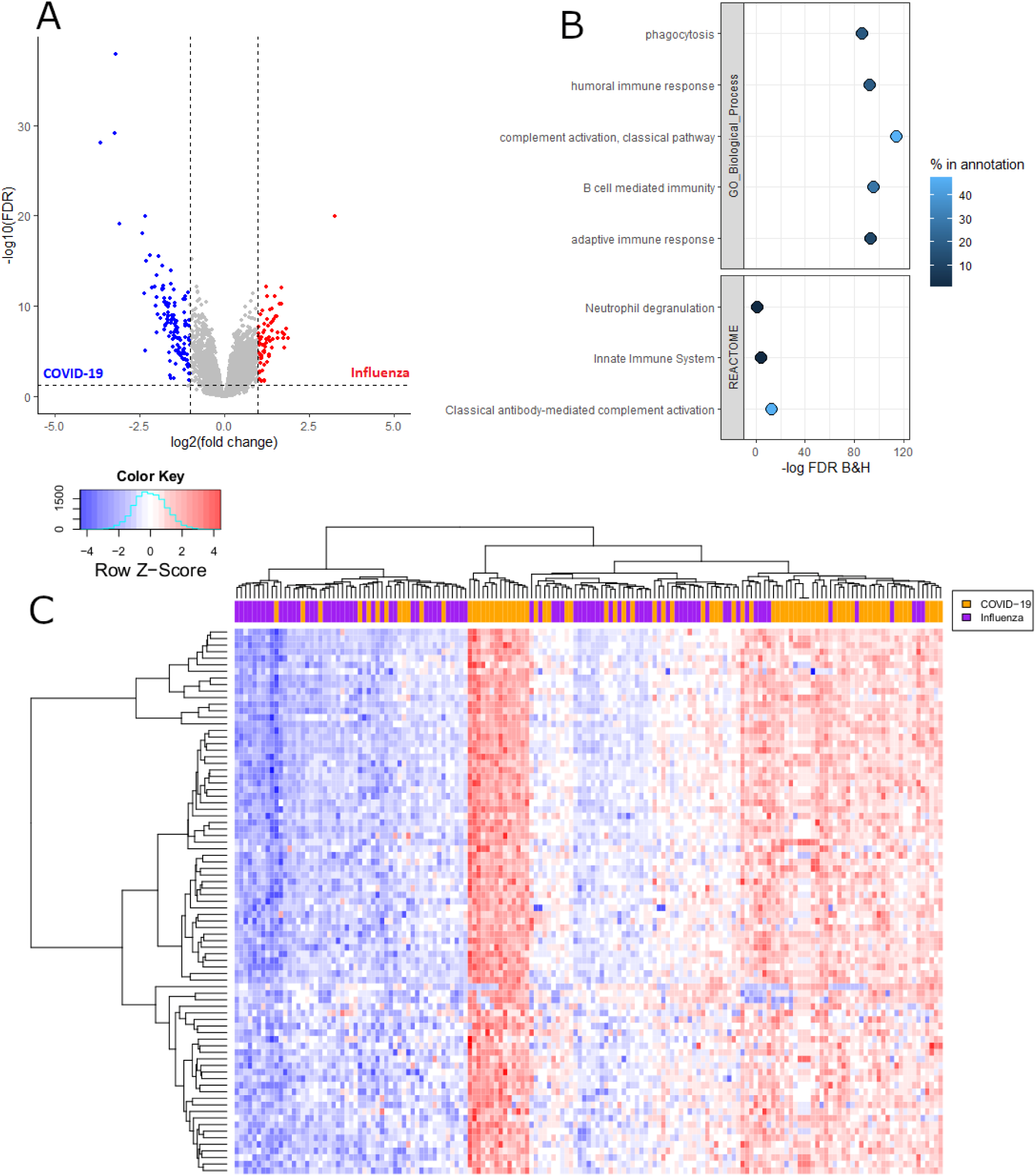
Increased adaptive immune response in patients with COVID-19 compared to influenza. A) Volcano plot of transcripts falling within the threshold values (FDR < 0.05 and log2 fold change < −1 or >1) which were used for enrichment analysis with ToppGene. B) Enrichment analysis of the transcripts with an increased abundance in patients with COVID-19 identified an increased adaptive immune response. Percentage in annotation is the ratio of the input query genes overlapping with the genes in the pathway database. C) Heatmap of 83 immunoglobulin gene transcripts, which overlap with the GO biological process term ‘adaptive immune response’, found at a higher abundance in patients with COVID-19. Positive Z-scores are seen mostly in patients with COVID-19 while negative Z-scores are mostly seen in patients with influenza.

83 immunoglobulin genes, associated with the GO biological process term ‘adaptive immune response’, were found to have higher transcript abundance in the majority of patients with COVID-19 than those with influenza (p-value < 2.22×10^−16^, Wilcoxon test) (**Figure 3C** and **Supplementary figure 4**) and by using a total Z-score, patients with COVID-19 or influenza were classified as having either a high or low abundance of these 83 immunoglobulin genes. A high abundance was associated with a total positive Z-score (1.46 to 175.46) which was identified in 59 patients with COVID-19 and 21 patients with influenza indicating a higher than average abundance of these 83 adaptive immune response related immunoglobulin genes. While a low abundance was associated with a total negative Z-score (−0.12 to −154.93) identified in 19 patients with COVID-19 and 62 patients with influenza indicating a lower than average abundance of adaptive immune response related immunoglobulin genes. COVID-19 patients with lower abundance of adaptive immune response related immunoglobulin genes, a total negative Z-score, were found to be significantly older (p-value 6.32×10^−3^, T-test) and had a shorter duration of symptoms before being admitted into hospital (p-value 5.9×10^−04^, Wilcoxon test). Additionally, COVID-19 patients with high abundance of adaptive immune response related immunoglobulin genes, a total positive Z-score, were significantly more likely to be still alive 30 days after admitted into hospital (x^2^ 13.39 and p-value 2.52×10^−04^, Chi-square test) (**Figure 4**).

**Figure 4:**
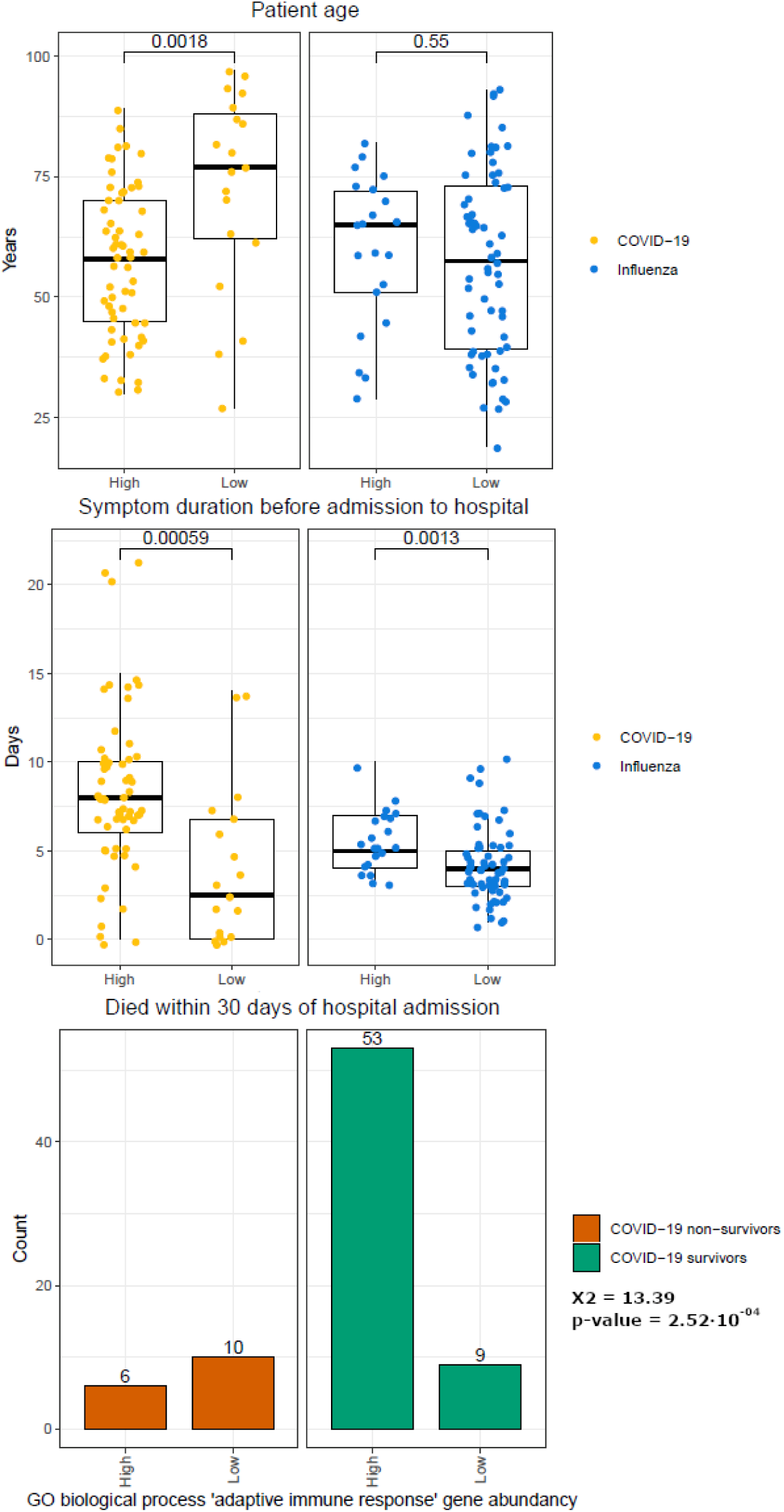
Comparison of 83 immunoglobulin gene abundances in patients with COVID-19 or influenza. Metadata of patients with COVID-19 or influenza with a low or high abundance of the 83 immunoglobulin genes related to GO biological process term ‘adaptive immune response’ were compared. Significant differences were detected based on age (patients with COVID-19, p-value 1.8×10^−3^, Wilcoxon-test) and duration of symptoms (patients with COVID-19, p-value 5.9×10^−04^ and patients with influenza, 1.3×10^−03^, Wilcoxon test), with older individuals and shorter symptom duration associated with the low immunoglobulin gene abundance group for COVID-19. Additionally, a low abundance of immunoglobulin genes was associated with decreased COVID-19 survival (x^2^ 13.39 and p-value 2.52×10^−04^, Chi-square test).

### Topological mapping of global gene patterns

Topological analysis allows the measurement of the global profiles of transcript abundances relative to gene pathways without data reduction and this was used to define a global map of differentially activated pathways between COVID-19 and influenza. The first differentially activated TopMD pathway was enriched for ribosomal and insulin related pathways, with peak gene *UBA52*: named by GO analysis as cytoplasmic ribosomal proteins (adjusted p-value 1.55×10^−146^). This pathway was also found to be enriched for genes expressed by transcription factor Myc (adjusted p-value 7.07×10^−53^) against the ChEA 2016 transcription factor database and of dendritic cells in the ARCHS4 transcription factors’ co-expression database (adjusted p-value 1.34×10^−36^). Activated Myc represses interferon regulatory factor 7 (*IRF7*) and a significant lower abundance of *IRF7* was found in patients with COVID-19 compared to influenza (**Supplementary figure 5**). The second differentially activated TopMD pathway had peak gene *NDUFAB1*; named by GO analysis as mitochondrial complex I assembly model OXPHOS system WP4324 (adjusted p-value 2.81×10^−66^). The third differentially activated TopMD pathway was named by GO analysis as proteasome degradation WP183 (adjusted p-value, 1.46×10^−64^), with *PSMD14* as the peak gene (**Figure 5** with full detail in **Additional file 3** and the global map of differentially activated pathways available online (65)).

**Figure 5:**
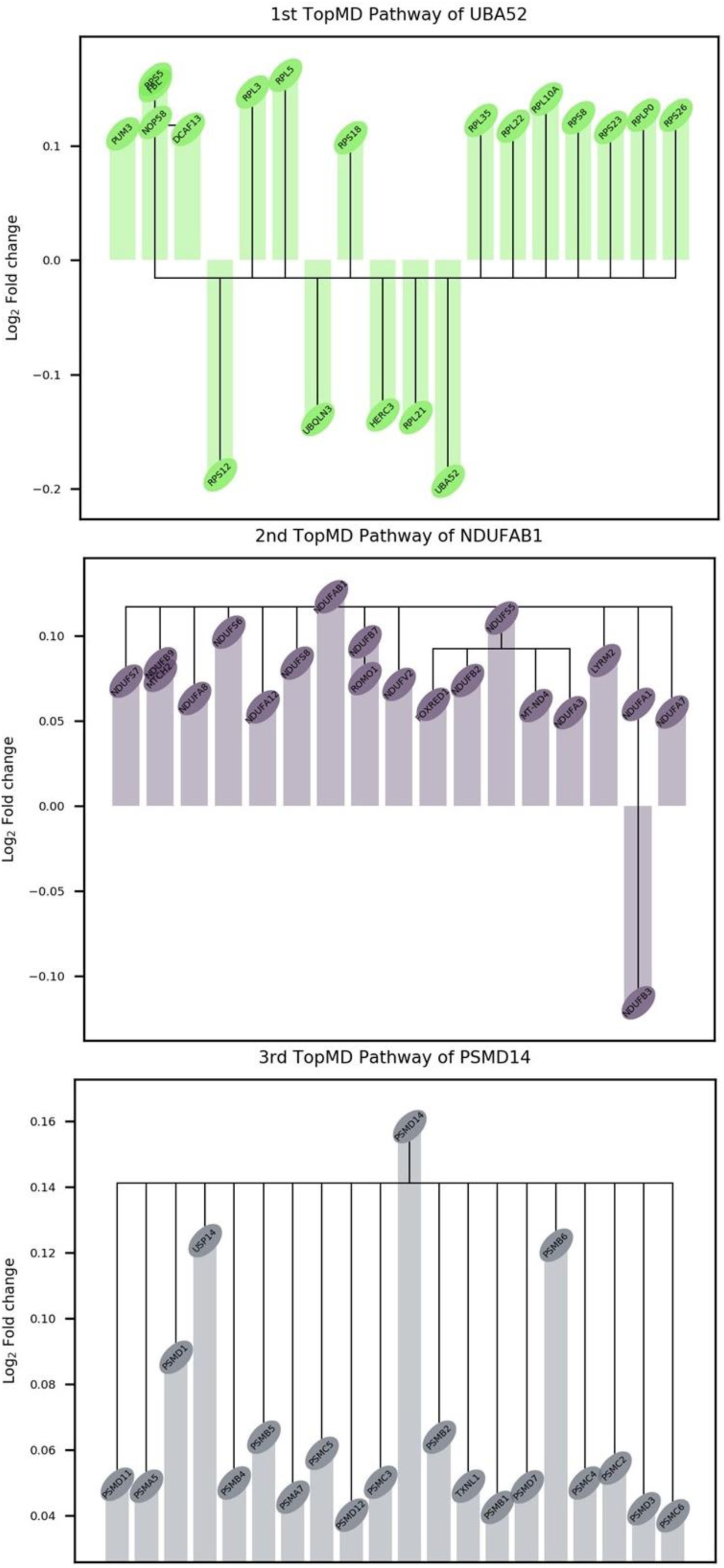
Differentially activated pathways between hospitalised patients with COVID-19 or influenza identified with topological analysis. The difference (Log2 fold change) in patients with COVID-19 compared to patients with influenza is plotted for the top 20 genes of the 1^st^, 2^nd^ and 3^rd^ TopMD pathways.

### Cell subsets supporting innate and adaptive immune response differences

Analysis of the blood transcriptome can be used to predict the immune cells present (64). Levels of different predicted cell types were assessed to determine whether there were differences in immune system associated cells between patients with COVID-19 or influenza (**Figure 6**). Statistical testing was done on cell type levels identified with CIBERSORTx. M0 macrophages (p-value 3.63×10^−06^), plasma cells (p-value 5.05×10^−04^), cytotoxic CD8+ T cells (p-value 4.58×10^−03^), regulatory T cells (p-value 7.30×10^−03^) and resting natural killer cell (p-value 8.90×10^−03^) were found to be significantly higher in COVID-19 patients, while in influenza patients activated dendritic cells (p-value 2.23×10^−02^) were significantly higher.

**Figure 6:**
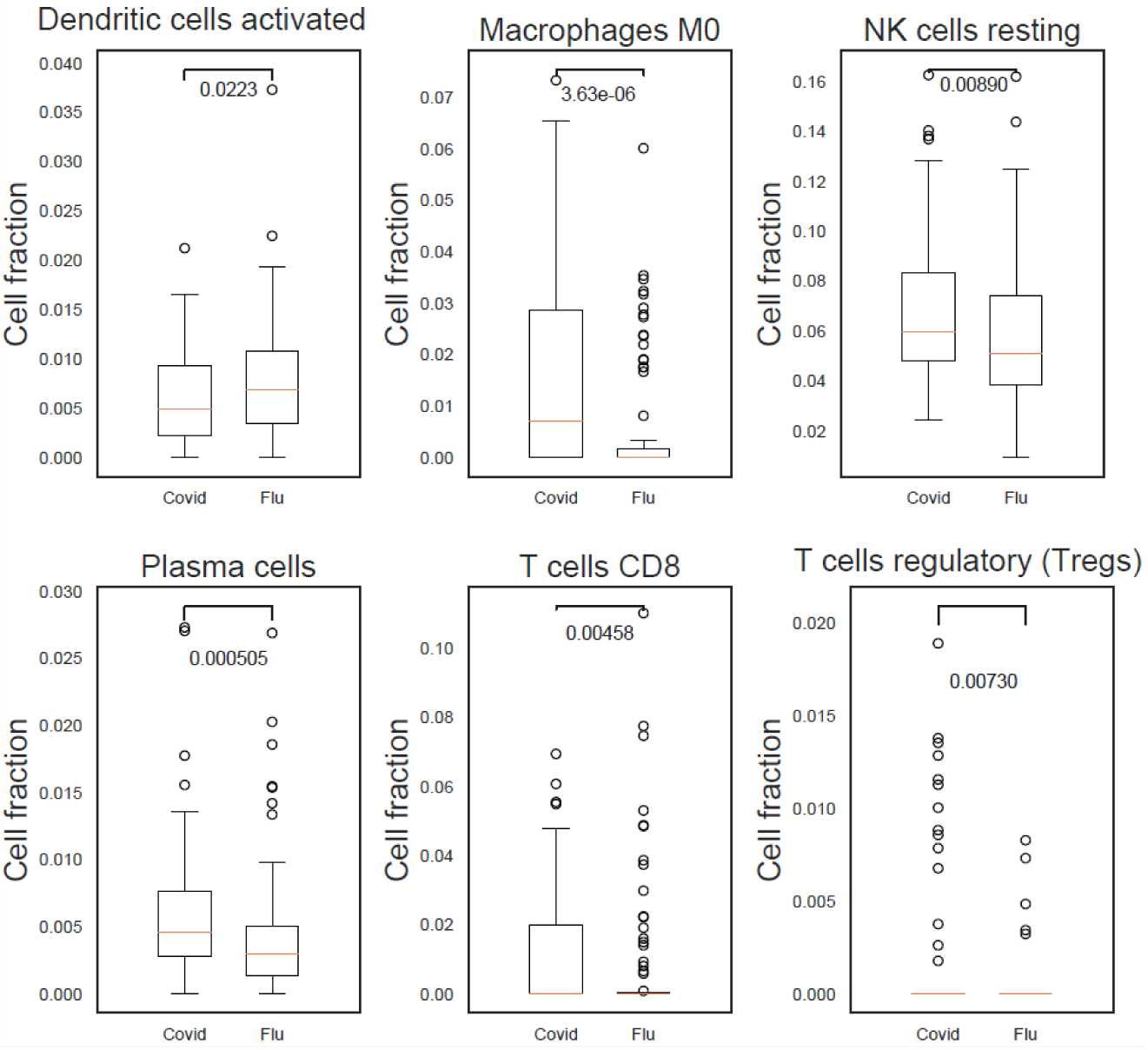
Increase of predicted cells associated with an innate and adaptive immune response in COVID-19 patients. M0 macrophages, plasma cells, cytotoxic CD8+ T cells, regulatory T cells and resting natural killer (NK) cells were found to be significantly higher in COVID-19 patients. In influenza patients a significantly higher proportion of activated dendritic cells was detected.

Predicted cell type levels between COVID-19 survivors and non-survivors indicated an increase of neutrophils (p-value 2.84×10^−04^) in patients who died of COVID-19. In contrast, an increase of naïve CD4+ T cells (p-value 1.92×10^−03^), M0 macrophages (p-value 1.20×10^−02^), M2 macrophages (p-value 1.48×10^−02^), naïve B cells (p-value 1.57×10^−02^) and naïve cytotoxic CD8+ T cells (p-value 2.31×10^−02^), were identified in patients who went on to survive COVID-19 (**Figure 7**).

**Figure 7:**
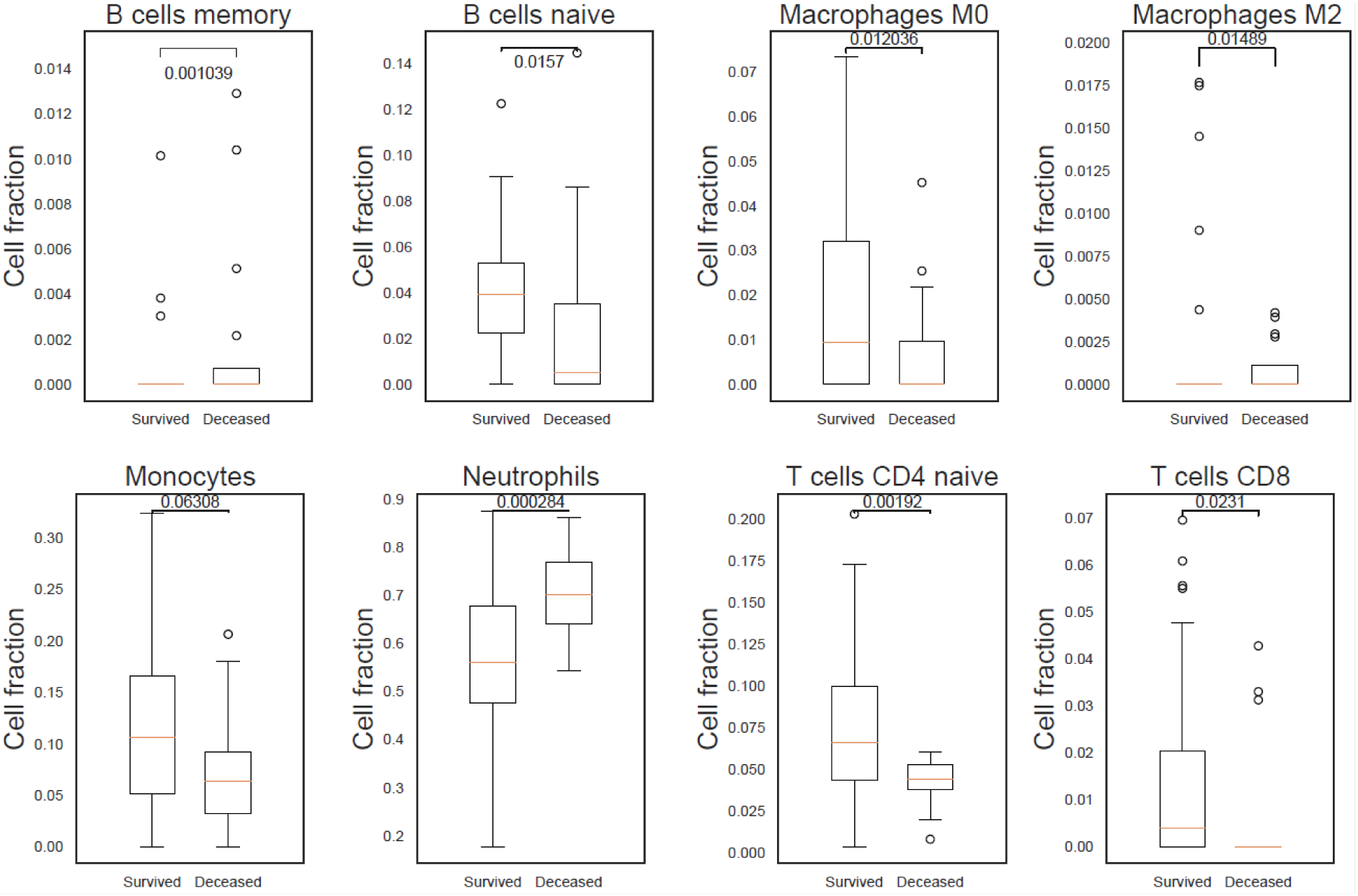
Differences in immune response indicated by predicted cell types in COVID-19 survivors and non-survivors. A) A statistically significant higher count of neutrophils in COVID-19 patients who died after 30 days indicating the presence of an elevated innate immune response. B) Adaptive immune response in COVID-19 survivors as can be seen by the statistically significant higher count of naïve B cells, and CD4+ and CD8+ T cells.

### Efficient adaptive immune response associates with COVID-19 survival

As already noted, a high abundance of the GO biological process ‘adaptive immune response’ related transcripts, mostly immunoglobulin genes, was associated with COVID-19 survival (**Figure 4**). Here a direct assessment was done of the blood transcriptome differences between patients who, at 30 days after hospital admission, survived or who died of COVID-19. A total of 23,850 abundance measures of gene transcripts were obtained after filtering out transcripts with low counts, of which 6.645 transcripts were found to be significant (FDR p-value < 0.05) of which, 537 transcripts exceeded a log2 fold change of < −1 or > 1, with 265 transcripts showing higher abundance in patients who survived COVID-19 and 272 transcripts showing a higher abundance in patients who died of COVID-19 (**Figure 8A** and **Additional file 4**).

**Figure 8:**
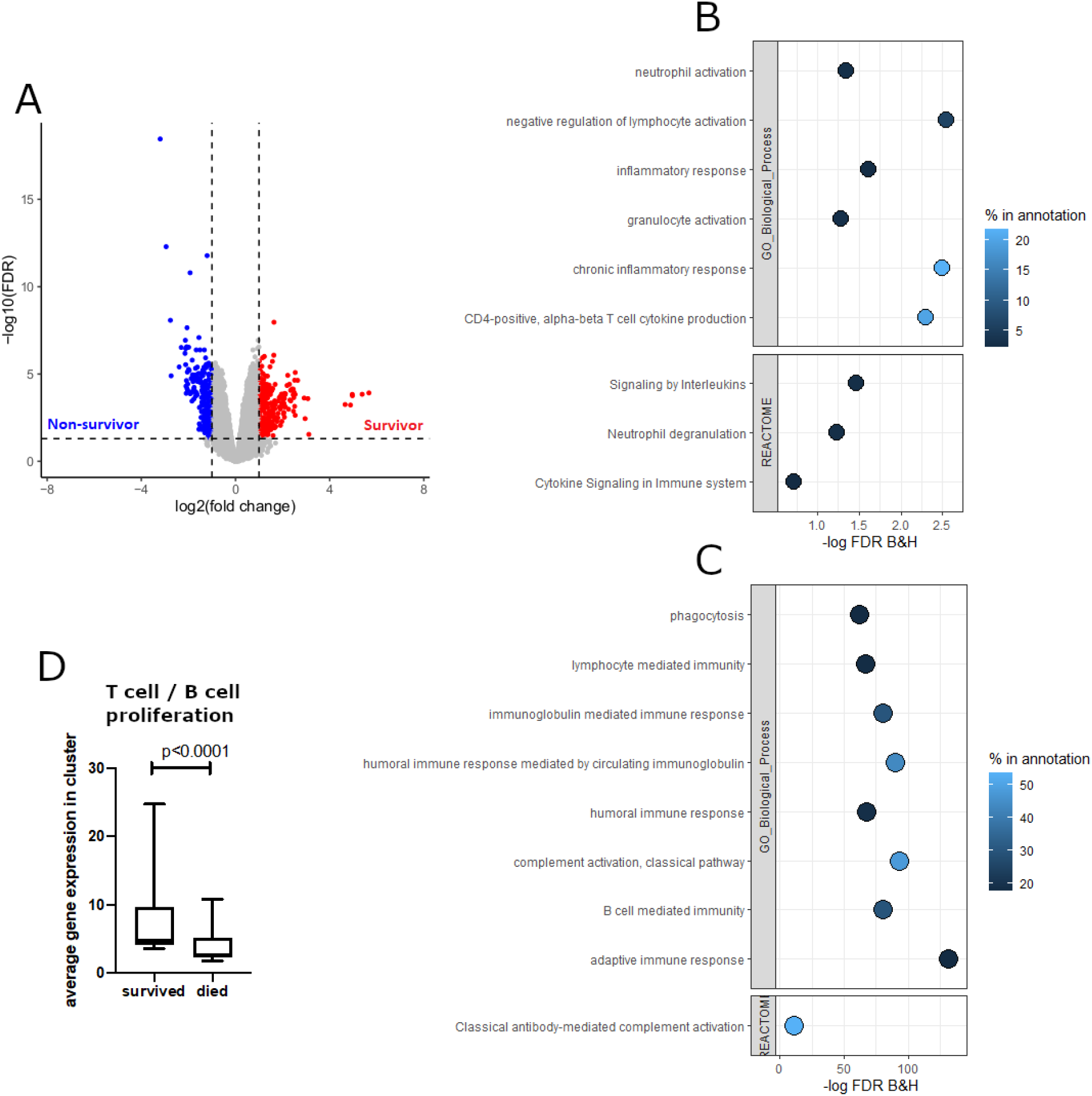
Increased innate immune response in COVID-19 non-survivors and increased adaptive immune response in COVID-19 survivors. A) Volcano plot of transcripts falling within the threshold values (FDR < 0.05 and log fold change < −1 or >1) which were used for enrichment analysis with ToppGene. B) Enrichment analysis identified an increased innate immune response in patients who died of COVID-19 after 30 days of hospital admission. Percentage in annotation is the ratio of the input query genes overlapping with the genes in the pathway database. C) An enrichment of adaptive immune response related pathways was detected in patients with COVID-19 who were still alive 30 days after hospital admission. Percentage in annotation is the ratio of the input query genes overlapping with the genes in the pathway database. D) Increase of T cell and B cell proliferation in COVID-19 survivors (paired non-parametric T-test).

In patients who died of COVID-19 an enrichment for biological processes involved with an inflammatory response including interleukin signalling and neutrophil activation and degranulation was detected (**Figure 8B**). While in COVID-19 survivors biological processes involved with the adaptive immune system including complement activation, B cell mediated immunity and a humoral immune response mediated by circulating immunoglobulins was found to be enriched (**Figure 8C**). Additionally, transcript abundances associated with T cell and B cell proliferation were significantly higher in COVID-19 survivors (p-value < 1.0×10^−04^, paired non-parametric T-test) (**Figure 8D**).

### Immune signatures as predictors of COVID-19 outcome

Distinct immune signature genes were selected and assessed for their prediction accuracy in stratifying patients with COVID-19 for disease outcome, fatality or survival. A signature of 47 genes was identified (**Figure 9A**), representative of the four biggest clusters of genes associated with either patients with COVID-19 who survived or died. The associated GO biological process terms were ‘humoral immune response mediated by circulating immunoglobulin’ (FDR p-value 2.23×10^−46^), ‘nucleosome assembly’ (FDR p-value 5.46×10^−19^), ‘regulation of T-helper 1 cell cytokine production’ (FDR p-value 4.24×10^−03^) and ‘regulation of T cell activation’ (FDR p-value 4.51×10^−04^) (**Supplementary figure 6**). This was highly predictive for outcome, with a maximum specificity of 75% and sensitivity of 93% (**Figure 9B** and **Table 4**).

**Figure 9:**
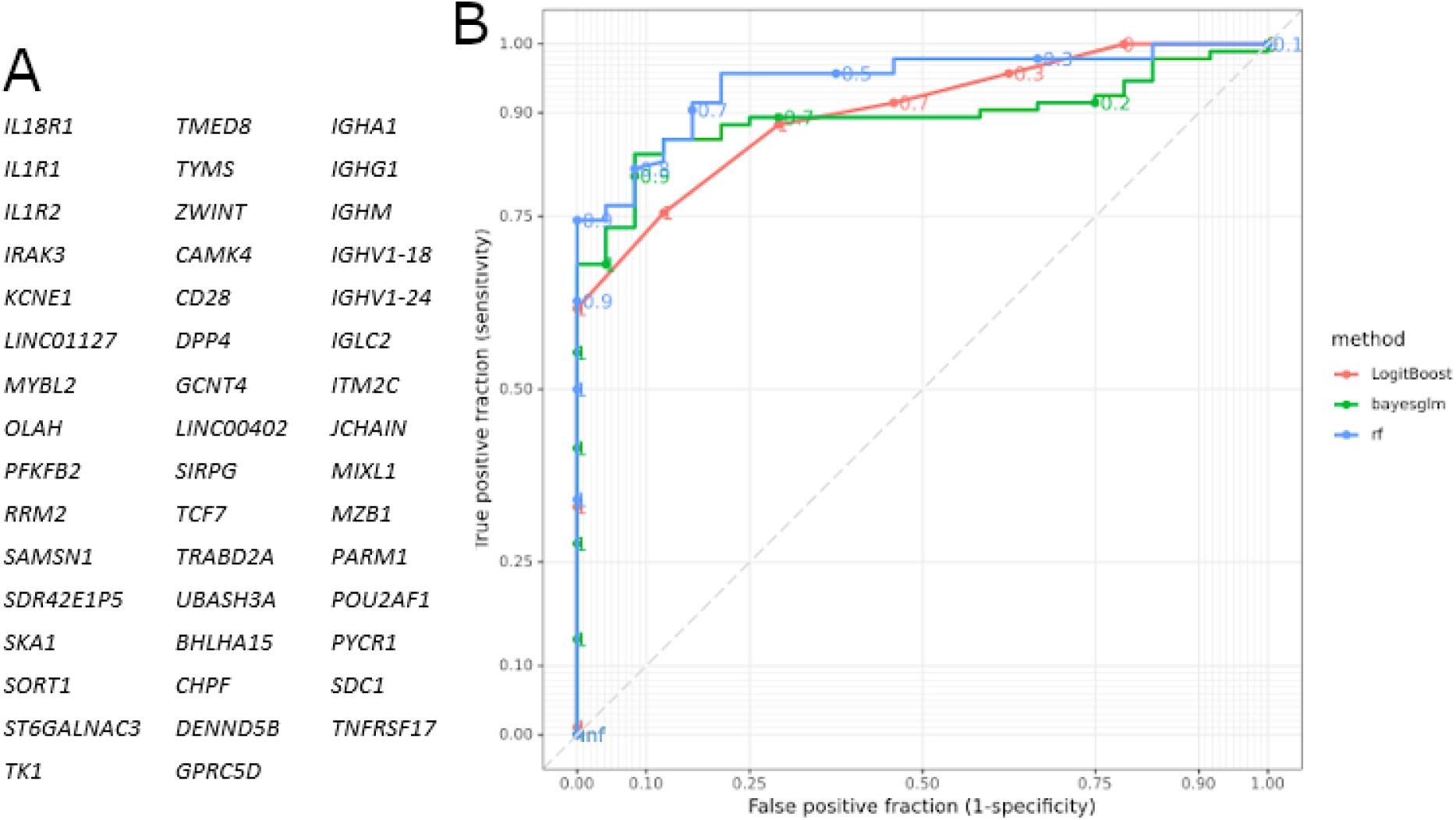
Receiver Operating Characteristic (ROC) curves showing prediction accuracy COVID-19 survivors and non-survivors. A) Genes identified with EdgeR and gene co-expression analysis and used for subsequent modelling. B) ROC curves according to the three models used (Boosted Logistic Regression (LogitBoost), Bayesian Generalised Linear (Bayesglm) and RandomForest (rf)).

**Table 4:** Potential for stratifying patients with COVID-19 upon admission to hospital on likely disease outcome.

| ModelName | Accuracy | TrainAUC | PredictAUC | Sensitivity | Specificity |
| --- | --- | --- | --- | --- | --- |
| rf | 0.7368 | 0.96 | 0.9 | 0 | 0.9333 |
| LogitBoost | 0.8421 | 0.9381 | 0.875 | 0.75 | 0.8667 |
| Bayesglm | 0.8421 | 0.895 | 0.7333 | 0.25 | 1 |

In total three different models were used (RandomForest (rf), Boosted Logistic Regression (LogitBoost) and Bayesian Generalised Linear (Bayesglm)). The 47 genes identified with gene co-expression and differential gene expression analysis were used as input. The highest sensitivity obtained was 75% and for specificity 93%.

## Discussion

As previously reported transcriptomic analysis of blood samples provide a relatively non-invasive window on the immune response, as previously shown by differentiating patients with Ebola virus disease at the acute stage (66). In this study we explored the functional blood transcriptomic differences, focussing mainly on the immune response, between patients with COVID-19, admitted to hospital during the first wave of the pandemic, and patients with a well-characterised stereotypical seasonal respiratory virus infection, influenza. Furthermore, we compared the blood transcriptomes of COVID-19 survivors and non-survivors for promising prognostic signatures indicative of COVID-19 survival.

35 variables that can provide prognostic information on COVID-19 associated mortality and 14 variables that can provide prognostic information on COVID-19 severity have previously been reported (67). We compared these known COVID-19 prognostic variables (67) between patients with COVID-19 or influenza and found more active smokers among influenza patients. High C-reactive protein (CRP), which previously has been reported to be similar upon admission to hospital between patients with COVID-19 or influenza (3), hypertension and diabetes were more common among patients with COVID-19. We also found an increase of liver disease, which has been classified as a low or very low certainty predictor (67), in patients with COVID-19. In our cohort more patients with influenza had an underlying respiratory disease. Similar to what has been previously reported (3) upon admission to hospital both patients with COVID-19 or influenza presented with similar WBC and neutrophil counts, and although we detected a difference in lymphocytes between patients with COVID-19 or influenza, there was no difference the N/L ratio. Similar to Piroth *et al.* (4) we found that the average length of stay was higher for patients with COVID-19 compared to influenza, and more patients with COVID-19 needed supplementary oxygen, and finally while Piroth *et al.* (4) report a roughly three times higher relative risk of death for COVID-19, in our cohort no influenza patients died whilst admitted to hospital and so this could not be assessed. In addition, we compared COVID-19 survivors and non-survivors, and as reported the high certainty prognostic variables for mortality and/or severity of increased age, hypertension, cardiovascular disease, diabetes, underlying respiratory disease (including COPD) and a high WBC (67) were increased in non-survivors. While it has previously been reported that CRP and N/L ratio were elevated in patients with COVID-19 who became critically ill (3), in our study we saw no difference in CRP, neutrophil count and lymphocyte count between COVID-19 survivors and non-survivors. However, we found that urea, creatinine, alanine aminotransferase, troponin and several cytokines, including IL-1β, IL-6, IL-8, IL-10 and TNFα, to be higher in patients who died of COVID-19.

Our initial global analysis of blood transcriptomic differences between patients with COVID-19 or influenza detected contrasting innate and adaptive immune programmes. An impaired immune response to viruses and interferon signalling in patients with COVID-19 was found, as described previously (6–9), compared to patients with influenza, which are known to produce an IFN response (3). Furthermore, in accordance with accumulating evidence of aberrant blood clotting in patients with COVID-19 (68, 69), transcripts expressed by megakaryocytes and platelets associated with blood coagulation were in a higher abundance in COVID-19 patients. Gene clusters associated with an innate immune response were found to be associated with influenza. While, in contrast, gene clusters associated with an adaptive immune response and an increase of predicted plasma cells and CD8+ T cells with COVID-19, pointing to T cell and B cell activation / priming.

Further analysis revealed various immunoglobulin genes had increased transcript abundance in patients with COVID-19 compared to patients with influenza. This significant over representation of a wide range of heavy chain and light chain V genes in patients with COVID-19 has been described before (70) and the implementation of antibody analysis in plasma samples has been used as an additional tool in diagnosing COVID-19 (71). We found that the 86.8% (53/61) of patients who survived COVID-19 had a higher than average transcript abundance of 83 immunoglobulin genes, which overlap with the GO biological term ‘adaptive immune response’, while this was 37.5% among the patients who died of COVID-19. Further analysis revealed that the aforementioned higher than average transcript abundance is associated with a younger age of the patient, a longer symptom duration before admittance into hospital and a positive survival outcome 30 days after hospital admission. A lower than average transcript abundance of 83 immunoglobulin genes was detected in 62.5% (10/16) of patients who died of COVID-19, compared to 14.8% (9/61) of patients who survived COVID-19.

We subsequently detected an increased transcript abundance from genes associated with T cell and B cell proliferation, an enrichment for gene pathways involved with an adaptive immune response, and an increase in predicted CD4+ and CD8+ T cells and naïve B cells in patients who survived COVID-19, highlighting the importance of an efficient adaptive immune response as previously reported (17). The predicted cell fraction of naïve CD4+ T cell was found to be higher compared to CD8+ T cells indicating a higher CD4+ T cell response to SARS-CoV-2 than a CD8+ T cell response, supporting previous observations (17, 72), which has been found to control primary SARS-CoV-2 infection (22). We note that the CD8+ T cells were mostly seen in COVID-19 survivors, compared to COVID-19 non-survivors, which has been associated with a positive COVID-19 outcome (22, 73).

In contrast, we detected in COVID-19 non-survivors an enrichment of pathways involved with the negative regulation of lymphocyte activation and increased neutrophil activation and degranulation, supported by a significant decrease in predicted cell fraction of naïve B cells and naïve CD4+ and CD8+ T cells and an increase of the neutrophil cell fraction. This is consistent with previous studies finding elevated levels of neutrophils in blood (74) and lungs (75–78) in severe COVID-19. Furthermore, gene pathways involved with an inflammatory response and cytokine signalling were enriched in COVID-19 non-survivors and we detected that a higher transcript abundance of several IL genes (*IL1-RAP*, *IL-10*, *IL1-R1*, *IL1-R2*, *IL18-R1* and *IL18-RAP*) and laboratory results indicated a increase of TNFα, IL-1β, IL-8, and IL-33 with the largest increase for IL-6 and IL-10. This is consistent with the previously reported positive regulation of genes encoding the activation of innate immune system, viral and IFN response (3), and increase of proinflammatory macrophages (79) and elevated IL-6 and IL-10 in severe COVID-19 cases (14–16).

When comparing the immune response between patients who either survived or died of COVID-19 it appears that, as Sette and Crotty (24) summarised, that COVID-19 severity is largely due to an early virus-driven evasion of innate immune recognition leading to a subsequent delayed adaptive immune response with a fatal COVID-19 outcome, as shown by Lucas *et al.* (80), where the innate immune response is ever-expanding due to an absence of a quick T cell response. A delayed adaptive immune response to COVID-19 can occur in the elderly due to their reduced ability to mount a successful adaptive immune response leading to an increased risk of death (22). This reduced ability to mount an adaptive immune response in the elderly is due to a scarcity of naïve T cells caused by aging (18–20) and the association of age and severe or fatal COVID-19 is already known, for example, as of April 15^th^ 2021 in the United States 95.4% of COVID-19 deaths have occurred in 50-year-olds and older, and 59.3% in 75-year-olds and older (81). Similarly, we found that patients who survived COVID-19 were younger, had a higher predicted naïve CD4+ T cell and naïve B cell fraction, and had an increased heart rate compared to non-survivors. Further research is needed to assess the causality of these factors, for example the relationship between increased age and heart rate in non-survivors.

Topological analysis was performed to identify the global map of gene pathways differentially activated between COVID-19 and influenza. The first differentially activated pathway was enriched for genes related to ribosomal and insulin pathways indicating differences in effects on translational machinery and supporting the reported roles of insulin resistance linked to COVID-19 severity (82). Although highly speculative, insulin signalling differences may reflect the role of angiotensin converting enzyme 2 (ACE2), the binding site for SARS-CoV-2, which degrades angiotensin 2, protecting against oxidative stress and insulin resistance driven by the renin-angiotensin-aldosterone system (83). Additionally, ACE2 expression has been found to be increased in rats given a high sucrose diet or insulin sensitisers (84). Furthermore, the first pathway was also found to be enriched for genes transcribed by Myc. Activated Myc represses *IRF7* which regulates type I IFN production (85), and correspondingly we found a significant lower *IRF7* expression and a lower induction of IFN in patients with COVID-19 compared to influenza. This low IFN induction in COVID-19 may be due to the virus avoiding or delaying an intracellular innate immune response to type I and type III IFNs (6–9). The second most differentially activated pathway, peak gene *NDUFAB1,* involved with the mitochondrial complex I assembly model OXPHOS system supports reported increased COVID-19 disease severity linked to SARS-CoV-2 being able to highjack mitochondria of immune cells, replicate and disrupt mitochondrial dynamics (86). The third differentially activated pathway was associated with the cellular ubiquitin-proteasome pathways which are known to play important roles in coronavirus infection cycles (87). The protein synthesis and ubiquitination-related pathways might reflect mechanisms of increased viral replication and suppression of host interferon signalling pathways, including increased degradation of IκBα which suppresses the IFN-induced NF-κB activation pathway. Also, in SARS-CoV, accessory protein P6, whose sequence is conserved in SARS-CoV-2 (88), promotes the ubiquitin-dependent proteasomal degradation of N-Myc interactor, thus limiting IFN signalling (89). However, the peak marker of this pathway *PSMD14* which prevents interferon regulatory factor 3 (*IRF3*) autophagic degradation and therefore, permits IRF3-mediated type I IFN activation (90); shedding light on the complex mechanistic differences regulating interferon production between COVID-19 and influenza.

## Conclusions

In this study, we have compared side-by-side SARS-CoV-2 and a stereotypical respiratory viral infection (influenza), and COVID-19 survivors and non-survivors. Distinct patterns of transcript abundances and cellular composition were found in whole blood that can differentiate the infection source, furthering our understanding of the antiviral immune response differences. Additionally, we observed a proinflammatory signature associated with a negative outcome in patients with COVID-19. Finally, a signature of transcript abundances in the blood transcriptome of COVID-19 patients, upon admission to hospital, was identified with prognostic potential to stratify patients into those likely to survive or die.

## Supporting information

Additional file 2: Differential gene expression results between patients with COVID-19 or influenza.

Additional file 3: Gene pathways identified by TopMD topological analysis between patients with COVID-19 or influenza.

Additional file 4: Differential gene expression results between COVID-19 survivors and non-survivors.

Additional file 5: Differential splicing results and discussion

Additional file 6: Differential splicing results between patients with COVID-19 or influenza.

## Data Availability

Following publication of major outputs all anonymised data will be made available on reasonable request to the corresponding author providing this meets local ethical and research governance criteria.

## Declarations

### Ethics approval and consent to participate

The COV-19POC trial was approved by the South Central - Hampshire A Research Ethics Committee (REC): REC reference 20/SC/0138 (March 16^th^, 2020); and REC reference 17/SC/0368 (September 7^th^, 2017) for the FluPOC trial. For full inclusion and exclusion criteria details see (25) and (26). Written informed consent was given by the patients, or consultee assent was obtained where patients were unable to give consent.

### Consent for publication

Not applicable.

### Authors’ contributions

TWC and DB conceptualized the study. SP and NJB screened and recruited the patients and collected the data in the FluPOC and CoV-19POC trials. RP-R and CH sample processing and experiments. J Legebeke, J Lord, RP-R, AFP, XD, FS, AG, JPRS, JAH and MEP performed data analysis. J Legebeke, J Lord, RP-R, AFP, FS, AG, JPRS, JAH and MEP drafted the article, and editing by J Legebeke, J Lord, RP-R, SP, NJB, GW, JPRS, PJS, JAH, MEP, TWC and DB. Project advice was given by JH, JSL, JAH, MP and TW. All authors read and approved the final manuscript.

## Acknowledgements

The authors would like to acknowledge and gives thanks to all the patients who kindly participated in the FluPOC and CoV-19POC studies and to all the clinical staff at University Hospital Southampton Foundation Trust who cared for them. In addition, the authors acknowledge the use of the IRIDIS High Performance Computing Facility, and associated support services at the University of Southampton, in the completion of this work.

## Supplementary information

### Supplementary figures

Additional file 1: Eighty-three immunoglobulin genes associated with the GO biological process term ‘adaptive immune response’ and their Z-scores.

Additional file 5: Differential splicing results and discussion

Additional file 6: Differential splicing results between patients with COVID-19 or influenza.

## References

1. ProMED. Promed Post [Internet]. Undiagnosed Pneumonia-China (Hubei): Request for information. 2019 [cited 2021 Jan 6]. Available from: https://promedmail.org/promed-post/?id=6864153%20#COVID19

2. World Health Organisation. WHO Coronavirus (COVID-19) Dashboard [Internet]. 2020 [cited 2021 May 1]. Available from: https://covid19.who.int

3. Galani I-E, Rovina N, Lampropoulou V, Triantafyllia V, Manioudaki M, Pavlos E, et al. Untuned antiviral immunity in COVID-19 revealed by temporal type I/III interferon patterns and flu comparison. Nat Immunol. 2021 Jan;22(1):32–40.

4. Piroth L, Cottenet J, Mariet A-S, Bonniaud P, Blot M, Tubert-Bitter P, et al. Comparison of the characteristics, morbidity, and mortality of COVID-19 and seasonal influenza: a nationwide, population-based retrospective cohort study. Lancet Respir Med. 2021 Mar;9(3):251–9.

5. Kreijtz JHCM, Fouchier R a. M, Rimmelzwaan GF. Immune responses to influenza virus infection. Virus Res. 2011 Dec;162(1–2):19–30.

6. Arunachalam PS, Wimmers F, Mok CKP, Perera RAPM, Scott M, Hagan T, et al. Systems biological assessment of immunity to mild versus severe COVID-19 infection in humans. Science. 2020 Sep 4;369(6508):1210–20.

7. Blanco-Melo D, Nilsson-Payant BE, Liu W-C, Uhl S, Hoagland D, Møller R, et al. Imbalanced Host Response to SARS-CoV-2 Drives Development of COVID-19. Cell. 2020 May 28;181(5):1036–1045.e9.

8. Laing AG, Lorenc A, Del Molino Del Barrio I, Das A, Fish M, Monin L, et al. A dynamic COVID-19 immune signature includes associations with poor prognosis. Nat Med. 2020 Oct;26(10):1623–35.

9. Bastard P, Rosen LB, Zhang Q, Michailidis E, Hoffmann H-H, Zhang Y, et al. Autoantibodies against type I IFNs in patients with life-threatening COVID-19. Science. 2020 Oct 23;370(6515).

10. Channappanavar R, Fehr AR, Vijay R, Mack M, Zhao J, Meyerholz DK, et al. Dysregulated Type I Interferon and Inflammatory Monocyte-Macrophage Responses Cause Lethal Pneumonia in SARS-CoV-Infected Mice. Cell Host Microbe. 2016 Feb 10;19(2):181–93.

11. Galani IE, Triantafyllia V, Eleminiadou E-E, Koltsida O, Stavropoulos A, Manioudaki M, et al. Interferon-λ Mediates Non-redundant Front-Line Antiviral Protection against Influenza Virus Infection without Compromising Host Fitness. Immunity. 2017 May 16;46(5):875–890.e6.

12. Monk PD, Marsden RJ, Tear VJ, Brookes J, Batten TN, Mankowski M, et al. Safety and efficacy of inhaled nebulised interferon beta-1a (SNG001) for treatment of SARS-CoV-2 infection: a randomised, double-blind, placebo-controlled, phase 2 trial. Lancet Respir Med. 2021 Feb 1;9(2):196–206.

13. Wang N, Zhan Y, Zhu L, Hou Z, Liu F, Song P, et al. Retrospective Multicenter Cohort Study Shows Early Interferon Therapy Is Associated with Favorable Clinical Responses in COVID-19 Patients. Cell Host Microbe. 2020 Sep 9;28(3):455–464.e2.

14. Chen G, Wu D, Guo W, Cao Y, Huang D, Wang H, et al. Clinical and immunological features of severe and moderate coronavirus disease 2019. J Clin Invest. 2020 May 1;130(5):2620–9.

15. Huang C, Wang Y, Li X, Ren L, Zhao J, Hu Y, et al. Clinical features of patients infected with 2019 novel coronavirus in Wuhan, China. Lancet Lond Engl. 2020 Feb 15;395(10223):497–506.

16. Liu T, Zhang J, Yang Y, Ma H, Li Z, Zhang J, et al. The role of interleukin-6 in monitoring severe case of coronavirus disease 2019. EMBO Mol Med. 2020 Jul 7;12(7):e12421.

17. Grifoni A, Weiskopf D, Ramirez SI, Mateus J, Dan JM, Moderbacher CR, et al. Targets of T Cell Responses to SARS-CoV-2 Coronavirus in Humans with COVID-19 Disease and Unexposed Individuals. Cell. 2020 Jun 25;181(7):1489–1501.e15.

18. Briceño O, Lissina A, Wanke K, Afonso G, Braun A von, Ragon K, et al. Reduced naïve CD8+ T-cell priming efficacy in elderly adults. Aging Cell. 2016;15(1):14–21.

19. Qi Q, Liu Y, Cheng Y, Glanville J, Zhang Z, Lee J-Y, et al. Diversity and clonal selection in the human T-cell repertoire. Proc Natl Acad Sci U S A. 2014 Aug 25;111(36):13139–44.

20. Wertheimer AM, Bennett MS, Park B, Uhrlaub JL, Martinez C, Pulko V, et al. Aging and cytomegalovirus infection differentially and jointly affect distinct circulating T cell subsets in humans. J Immunol. 2014 Mar 1;192(5):2143–55.

21. Morens DM, Fauci AS. Emerging Pandemic Diseases: How We Got to COVID-19. Cell. 2020 Sep 3;182(5):1077–92.

22. Moderbacher CR, Ramirez SI, Dan JM, Grifoni A, Hastie KM, Weiskopf D, et al. Antigen-Specific Adaptive Immunity to SARS-CoV-2 in Acute COVID-19 and Associations with Age and Disease Severity. Cell. 2020 Nov 12;183(4):996–1012.e19.

23. Magleby R, Westblade LF, Trzebucki A, Simon MS, Rajan M, Park J, et al. Impact of Severe Acute Respiratory Syndrome Coronavirus 2 Viral Load on Risk of Intubation and Mortality Among Hospitalized Patients With Coronavirus Disease 2019. Clin Infect Dis. 2020 Jun 30;(ciaa851).

24. Sette A, Crotty S. Adaptive immunity to SARS-CoV-2 and COVID-19. Cell. 2021 Feb 18;184(4):861–80.

25. Clark T. Evaluating the clinical impact of routine molecular point-of-care testing for COVID-19 in adults presenting to hospital: A prospective, interventional, non-randomised, controlled study (CoV19POC) [Internet]. 2020 [cited 2021 Mar 23]. Available from: https://eprints.soton.ac.uk/439309/2/CoV_19POC_Protocol_v2_0_eprints.pdf

26. Beard K, Brendish N, Malachira A, Mills S, Chan C, Poole S, et al. Pragmatic multicentre randomised controlled trial evaluating the impact of a routine molecular point-of-care ‘test-and-treat’ strategy for influenza in adults hospitalised with acute respiratory illness (FluPOC): trial protocol. BMJ Open. 2019 Dec 1;9(12):e031674.

27. Brendish NJ, Poole S, Naidu VV, Mansbridge CT, Norton NJ, Wheeler H, et al. Clinical impact of molecular point-of-care testing for suspected COVID-19 in hospital (COV-19POC): a prospective, interventional, non-randomised, controlled study. Lancet Respir Med. 2020 Dec;8(12):1192–200.

28. Clark TW, Beard KR, Brendish NJ, Malachira AK, Mills S, Chan C, et al. Clinical impact of a routine, molecular, point-of-care, test-and-treat strategy for influenza in adults admitted to hospital (FluPOC): a multicentre, open-label, randomised controlled trial. Lancet Respir Med. 2021 Apr 1;9(4):419–29.

29. Martin M. Cutadapt removes adapter sequences from high-throughput sequencing reads. EMBnet.journal. 2011 May 2;17(1):10–2.

30. Joshi NA, Fass JN. Sickle: A sliding-window, adaptive, quality-based trimming tool for FastQ files [Internet]. 2011. Available from: https://github.com/najoshi/sickl

31. Andrews S. FastQC: a quality control tool for high throughput sequence data. [Internet]. 2010. Available from: http://www.bioinformatics.babraham.ac.uk/projects/fastqc

32. Ewels P, Magnusson M, Lundin S, Käller M. MultiQC: summarize analysis results for multiple tools and samples in a single report. Bioinformatics. 2016 Oct 1;32(19):3047–8.

33. Dobin A, Gingeras TR. Mapping RNA-seq Reads with STAR. Curr Protoc Bioinforma. 2015;51(1):11.14.1–11.14.19.

34. Harrow J, Frankish A, Gonzalez JM, Tapanari E, Diekhans M, Kokocinski F, et al. GENCODE: the reference human genome annotation for The ENCODE Project. Genome Res. 2012 Sep;22(9):1760–74.

35. Shen S, Park JW, Lu Z, Lin L, Henry MD, Wu YN, et al. rMATS: Robust and flexible detection of differential alternative splicing from replicate RNA-Seq data. Proc Natl Acad Sci. 2014 Dec 23;111(51):E5593–601.

36. Li H, Handsaker B, Wysoker A, Fennell T, Ruan J, Homer N, et al. The Sequence Alignment/Map format and SAMtools. Bioinforma Oxf Engl. 2009 Aug 15;25(16):2078–9.

37. R Core Team. R: A Language and Environment for Statistical Computing [Internet]. 2020. Available from: https://cloud.r-project.org/index.html

38. RStudio Team. RStudio: Integrated Development Environment for R. [Internet]. 2020. Available from: http://www.rstudio.com/

39. Kassambara A. rstatix: Pipe-Friendly Framework for Basic Statistical Tests [Internet]. 2021 [cited 2021 Apr 23]. Available from: https://CRAN.R-project.org/package=rstatix

40. Rich B. table1: Tables of Descriptive Statistics in HTML [Internet]. 2021 [cited 2021 Apr 23]. Available from: https://CRAN.R-project.org/package=table1

41. Li S, Rouphael N, Duraisingham S, Romero-Steiner S, Presnell S, Davis C, et al. Molecular signatures of antibody responses derived from a systems biology study of five human vaccines. Nat Immunol. 2014 Feb;15(2):195–204.

42. Van Rossum G, Drake F. Python 3 Reference Manual. Scotts Valley, CA: CreateSpace; 2009.

43. Ritchie ME, Phipson B, Wu D, Hu Y, Law CW, Shi W, et al. limma powers differential expression analyses for RNA-sequencing and microarray studies. Nucleic Acids Res. 2015 Apr 20;43(7):e47.

44. Theocharidis A, Dongen S van, Enright AJ, Freeman TC. Network visualization and analysis of gene expression data using BioLayout Express 3D. Nat Protoc. 2009 Oct;4(10):1535–50.

45. Chen J, Bardes EE, Aronow BJ, Jegga AG. ToppGene Suite for gene list enrichment analysis and candidate gene prioritization. Nucleic Acids Res. 2009 Jul;37(Web Server issue):W305–311.

46. Anders S, Pyl PT, Huber W. HTSeq—a Python framework to work with high-throughput sequencing data. Bioinformatics. 2015 Jan 15;31(2):166–9.

47. Robinson MD, McCarthy DJ, Smyth GK. edgeR: a Bioconductor package for differential expression analysis of digital gene expression data. Bioinformatics. 2010 Jan 1;26(1):139–40.

48. Ashburner M, Ball CA, Blake JA, Botstein D, Butler H, Cherry JM, et al. Gene Ontology: tool for the unification of biology. Nat Genet. 2000 May;25(1):25–9.

49. Gene Ontology Consortium. The Gene Ontology resource: enriching a GOld mine. Nucleic Acids Res. 2021 Jan 8;49(D1):D325–34.

50. Wickham H. ggplot2: Elegant Graphics for Data Analysis [Internet]. 2016 [cited 2021 Apr 26]. Available from: https://ggplot2.tidyverse.org/authors.html

51. Kassambara A. ggpubr: “ggplot2” Based Publication Ready Plots [Internet]. 2020 [cited 2021 Apr 23]. Available from: https://CRAN.R-project.org/package=ggpubr

52. Strazzeri F, Schofield J, Skipp PJ, Sanchez-Garcia R, Koskela A, Sam M, et al. TopMD [Internet]. [cited 2021 May 1]. Available from: https://www.topmd.co.uk/

53. Franceschini A, Szklarczyk D, Frankild S, Kuhn M, Simonovic M, Roth A, et al. STRING v9.1: protein-protein interaction networks, with increased coverage and integration. Nucleic Acids Res. 2013 Jan;41(Database issue):D808–815.

54. Li YI, Knowles DA, Humphrey J, Barbeira AN, Dickinson SP, Im HK, et al. Annotation-free quantification of RNA splicing using LeafCutter. Nat Genet. 2018 Jan;50(1):151–8.

55. Knowles D, Li Y, Humphrey J, Pritchard J, Jenkinson G. Differential Splicing protocol [Internet]. [cited 2021 May 3]. Available from: https://davidaknowles.github.io/leafcutter/articles/Usage.html

56. LeafCutter Google group. Re: Leafcutter results [Internet]. [cited 2021 May 3]. Available from: https://groups.google.com/g/leafcutter-users/c/REkONdzrPfE/m/Jmm9rcLbBwAJ

57. Humphrey J, Knowles D, Li Y. LeafViz [Internet]. [cited 2021 May 3]. Available from: https://leafcutter.shinyapps.io/leafviz/

58. Vaquero-Garcia J, Barrera A, Gazzara MR, González-Vallinas J, Lahens NF, Hogenesch JB, et al. A new view of transcriptome complexity and regulation through the lens of local splicing variations. Valcárcel J, editor. eLife. 2016 Feb 1;5:e11752.

59. Newman AM, Steen CB, Liu CL, Gentles AJ, Chaudhuri AA, Scherer F, et al. Determining cell type abundance and expression from bulk tissues with digital cytometry. Nat Biotechnol. 2019 Jul;37(7):773–82.

60. Alizadeh lab, Newman lab. CIBERSORTx [Internet]. 2021 [cited 2021 May 3]. Available from: https://cibersortx.stanford.edu/

61. Tomic A, Tomic I, Waldron L, Geistlinger L, Kuhn M, Spreng RL, et al. SIMON: Open-Source Knowledge Discovery Platform. Patterns. 2021 Jan 8;2(1):100178.

62. Merkel D. Docker: lightweight Linux containers for consistent development and deployment. Linux J. 2014 Mar 1;2014(239):2:2.

63. Dorward DA, Russell CD, Um IH, Elshani M, Armstrong SD, Penrice-Randal R, et al. Tissue-Specific Immunopathology in Fatal COVID-19. Am J Respir Crit Care Med. 2021 Jan 15;203(2):192–201.

64. Liu X, Speranza E, Muñoz-Fontela C, Haldenby S, Rickett NY, Garcia-Dorival I, et al. Transcriptomic signatures differentiate survival from fatal outcomes in humans infected with Ebola virus. Genome Biol. 2017 Jan 19;18(1):4.

65. Strazzeri F, Schofield J, Skipp PJ, Sanchez-Garcia R, Koskela A, Sam M, et al. TopMD Global map between patients with COVID-19 or influenza [Internet]. [cited 2021 May 10]. Available from: https://topmd.co.uk/research/covidvflu

66. Bosworth A, Rickett NY, Dong X, Ng LFP, García-Dorival I, Matthews DA, et al. Analysis of an Ebola virus disease survivor whose host and viral markers were predictive of death indicates the effectiveness of medical countermeasures and supportive care. Genome Med. 2021 Jan 11;13(1):5.

67. Izcovich A, Ragusa MA, Tortosa F, Lavena Marzio MA, Agnoletti C, Bengolea A, et al. Prognostic factors for severity and mortality in patients infected with COVID-19: A systematic review. PloS One. 2020;15(11):e0241955.

68. Loo J, Spittle DA, Newnham M. COVID-19, immunothrombosis and venous thromboembolism: biological mechanisms. Thorax. 2021 Apr 1;76(4):412–20.

69. Eslamifar Z, Behzadifard M, Soleimani M, Behzadifard S. Coagulation abnormalities in SARS-CoV-2 infection: overexpression tissue factor. Thromb J. 2020 Dec 15;18(1):38.

70. Robbiani DF, Gaebler C, Muecksch F, Lorenzi JCC, Wang Z, Cho A, et al. Convergent antibody responses to SARS-CoV-2 in convalescent individuals. Nature. 2020 Aug;584(7821):437–42.

71. Guo L, Ren L, Yang S, Xiao M, Chang D, Yang F, et al. Profiling Early Humoral Response to Diagnose Novel Coronavirus Disease (COVID-19). Clin Infect Dis. 2020 Jul 28;71(15):778–85.

72. Sekine T, Perez-Potti A, Rivera-Ballesteros O, Strålin K, Gorin J-B, Olsson A, et al. Robust T Cell Immunity in Convalescent Individuals with Asymptomatic or Mild COVID-19. Cell. 2020 Oct 1;183(1):158–168.e14.

73. Peng Y, Mentzer AJ, Liu G, Yao X, Yin Z, Dong D, et al. Broad and strong memory CD4 + and CD8 + T cells induced by SARS-CoV-2 in UK convalescent individuals following COVID-19. Nat Immunol. 2020 Nov;21(11):1336–45.

74. Kuri-Cervantes L, Pampena MB, Meng W, Rosenfeld AM, Ittner CAG, Weisman AR, et al. Comprehensive mapping of immune perturbations associated with severe COVID-19. Sci Immunol [Internet]. 2020 Jul 15 [cited 2021 Apr 27];5(49). Available from: https://immunology.sciencemag.org/content/5/49/eabd7114

75. Li S, Jiang L, Li X, Lin F, Wang Y, Li B, et al. Clinical and pathological investigation of patients with severe COVID-19. JCI Insight. 2020 Jun 18;5(12).

76. Liao M, Liu Y, Yuan J, Wen Y, Xu G, Zhao J, et al. Single-cell landscape of bronchoalveolar immune cells in patients with COVID-19. Nat Med. 2020 Jun;26(6):842–4.

77. Schurink B, Roos E, Radonic T, Barbe E, Bouman CSC, de Boer HH, et al. Viral presence and immunopathology in patients with lethal COVID-19: a prospective autopsy cohort study. Lancet Microbe. 2020 Nov;1(7):e290–9.

78. Radermecker C, Detrembleur N, Guiot J, Cavalier E, Henket M, d’Emal C, et al. Neutrophil extracellular traps infiltrate the lung airway, interstitial, and vascular compartments in severe COVID-19. J Exp Med. 2020 Sep 14;217(2):1–11.

79. Schultze JL, Aschenbrenner AC. COVID-19 and the human innate immune system. Cell. 2021 Apr 1;184(7):1671–92.

80. Lucas C, Klein J, Sundaram ME, Liu F, Wong P, Silva J, et al. Delayed production of neutralizing antibodies correlates with fatal COVID-19. Nat Med. 2021 May 5;1–9.

81. Centers for Disease Control and Prevention. COVID Data Tracker [Internet]. Centers for Disease Control and Prevention. 2020 [cited 2021 Apr 15]. Available from: https://covid.cdc.gov/covid-data-tracker/index.html#demographics

82. Finucane FM, Davenport C. Coronavirus and Obesity: Could Insulin Resistance Mediate the Severity of Covid-19 Infection? Front Public Health. 2020;8(184):1–5.

83. Gheblawi M, Wang K, Viveiros A, Nguyen Q, Zhong J-C, Turner AJ, et al. Angiotensin-Converting Enzyme 2: SARS-CoV-2 Receptor and Regulator of the Renin-Angiotensin System: Celebrating the 20th Anniversary of the Discovery of ACE2. Circ Res. 2020 May 8;126(10):1456–74.

84. Takeda M, Yamamoto K, Takemura Y, Takeshita H, Hongyo K, Kawai T, et al. Loss of ACE2 exaggerates high-calorie diet-induced insulin resistance by reduction of GLUT4 in mice. Diabetes. 2013 Jan;62(1):223–33.

85. Honda K, Yanai H, Negishi H, Asagiri M, Sato M, Mizutani T, et al. IRF-7 is the master regulator of type-I interferon-dependent immune responses. Nature. 2005 Apr 7;434(7034):772–7.

86. Singh KK, Chaubey G, Chen JY, Suravajhala P. Decoding SARS-CoV-2 hijacking of host mitochondria in COVID-19 pathogenesis. Am J Physiol Cell Physiol. 2020 Aug 1;319(2):C258–67.

87. Raaben M, Posthuma CC, Verheije MH, te Lintelo EG, Kikkert M, Drijfhout JW, et al. The ubiquitin-proteasome system plays an important role during various stages of the coronavirus infection cycle. J Virol. 2010 Aug;84(15):7869–79.

88. Naqvi AAT, Fatima K, Mohammad T, Fatima U, Singh IK, Singh A, et al. Insights into SARS-CoV-2 genome, structure, evolution, pathogenesis and therapies: Structural genomics approach. Biochim Biophys Acta BBA - Mol Basis Dis. 2020 Oct 1;1866(10):165878.

89. Cheng W, Chen S, Li R, Chen Y, Wang M, Guo D. Severe acute respiratory syndrome coronavirus protein 6 mediates ubiquitin-dependent proteosomal degradation of N-Myc (and STAT) interactor. Virol Sin. 2015 Apr;30(2):153–61.

90. Wu Y, Jin S, Liu Q, Zhang Y, Ma L, Zhao Z, et al. Selective autophagy controls the stability of transcription factor IRF3 to balance type I interferon production and immune suppression. Autophagy. 2020 May 31;1–14.

