## Supplementary figures for "Distinct immune responses in patients infected with influenza or SARS-CoV-2, and in COVID-19 survivors, characterised by transcriptomic and cellular abundance differences in blood"

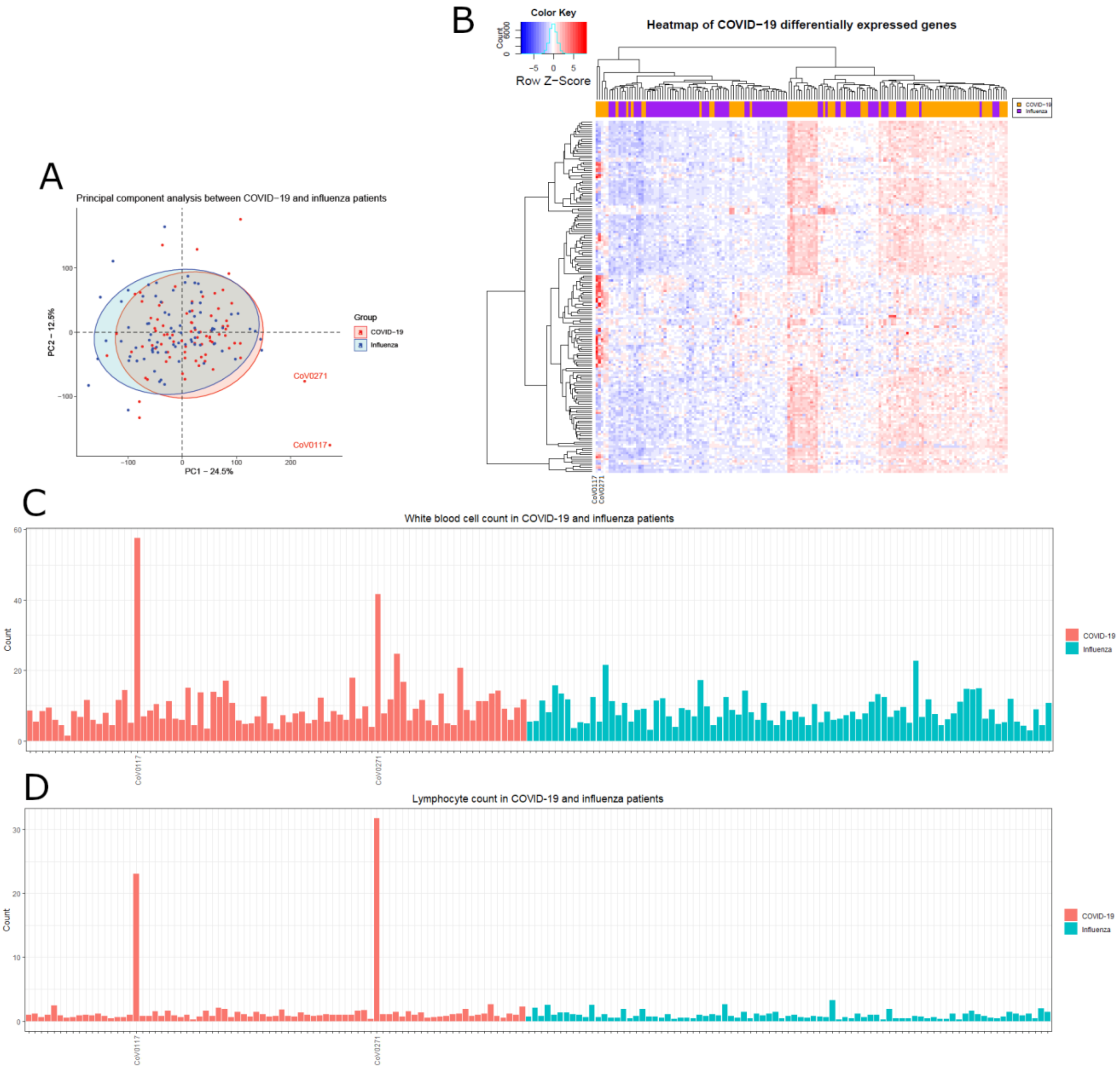


***Supplementary figure 1: Initial sample comparison revealed two COVID-19 patients which were subsequently excluded in further analyses.*** *A) Principal Component Analysis (PCA) graph comparing the COVID-19 patients with the influenza patients revealed two COVID-19 outliers (CoV0117 and CoV0271). B) Heatmap comparing the differentially expressed genes found specifically in COVID-19 patients indicates a different expression profile for CoV0117 and CoV0271 compared to the other samples. Assessment of the blood results indicates a high white blood cell count (C) and high lymphocyte count (D) in CoV0117 and CoV0271. Subsequent, analysis of the patient metadata revealed that these samples are from two patients with known chronic lymphocytic leukaemia.*

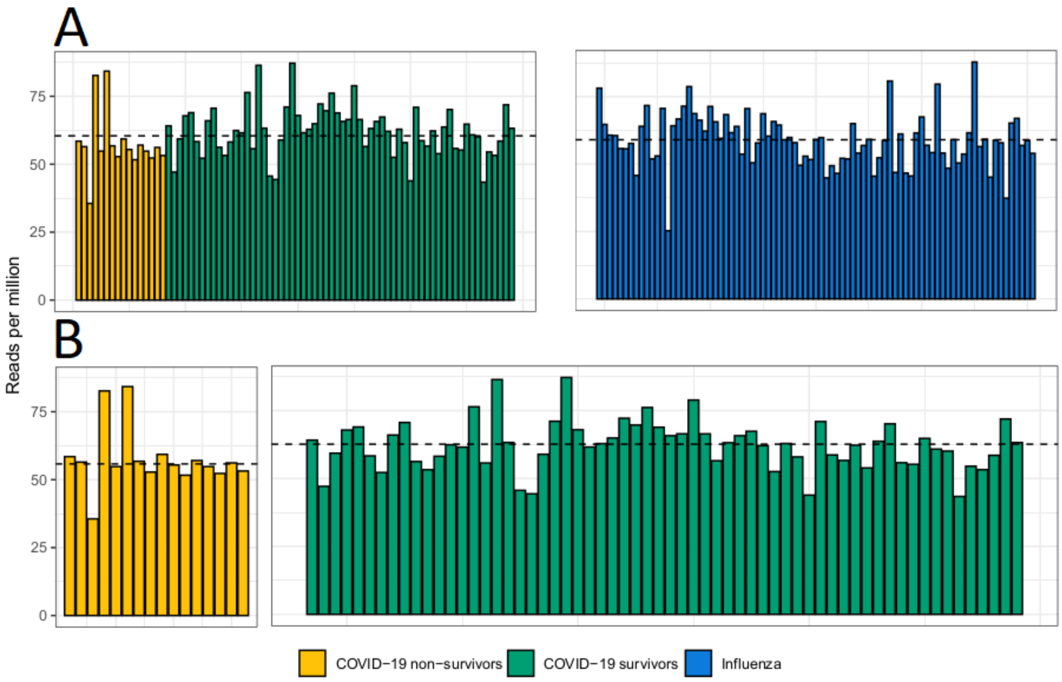


***Supplementary figure 2: Sequence read depths obtained in the patients with COVID-19 or influenza in reads per million.*** *A) Comparison of the sequence read depths obtained in the 78 patients with COVID-19 (median sequence depth of 60.4 million reads) and 83 patients with influenza (median sequence depth of 58.9 million reads). B) Sequence read depths in the patients with COVID-19 used for the comparison between survivors (median sequence depth of 62.6 million reads) and non-survivors (median sequence depth of 55.7 million reads). Median sequence depths are illustrated by the black dashed line.*

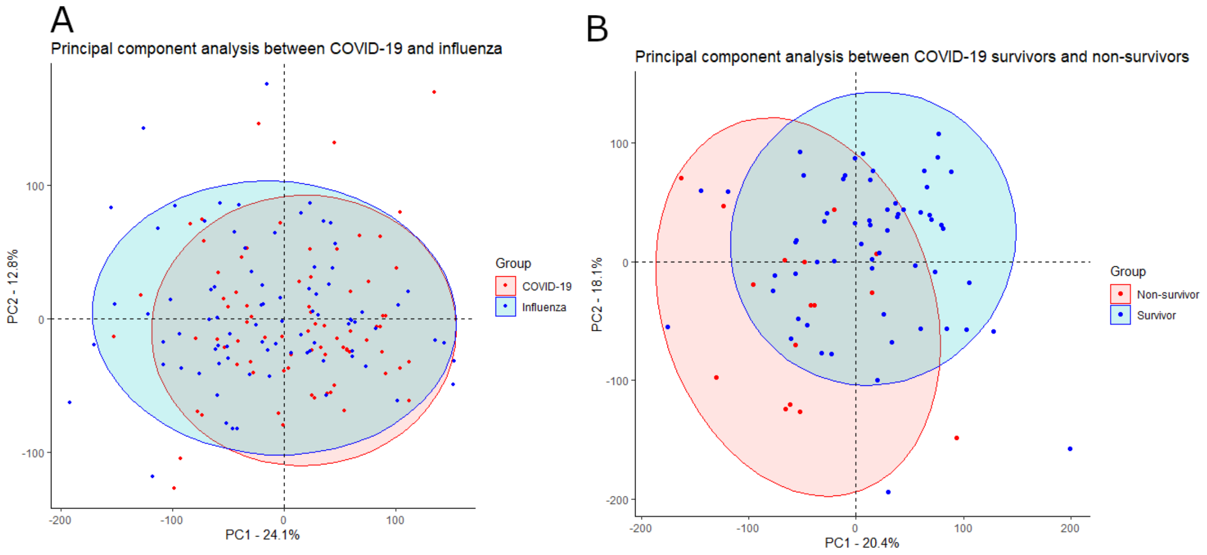


***Supplementary figure 3: PCA analysis between patients with COVID-19 or influenza, and who survived or died of COVID-19.*** *A) PCA analysis between patients with COVID-19 or influenza showing a substantial overlap between the two different group blood transcriptomes. B) A partial overlap of blood transcriptomes between patients who survived or who died of COVID-19.*

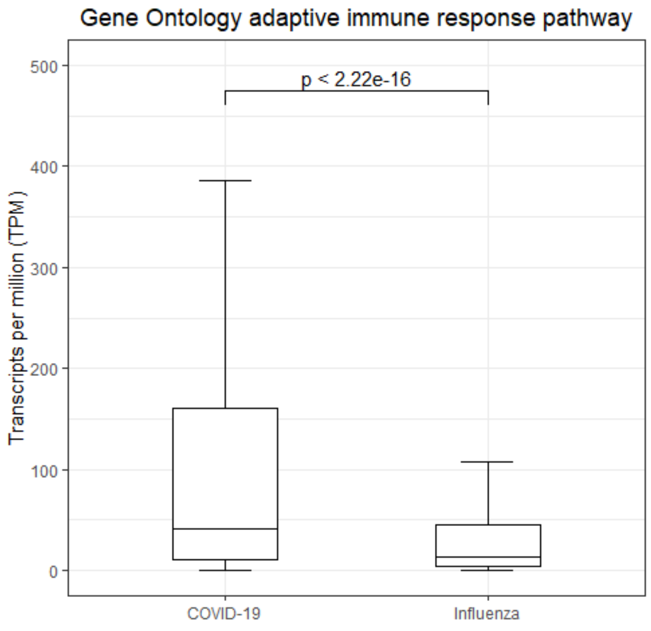


***Supplementary figure 4: GO Biological Process Term 'adaptive immune response' gene expression between patients with COVID-19 or influenza.*** *Unpaired non-parametric Wilcoxon test was used to assess the difference in abundancy of 83 immunoglobulin genes associated with the GO biological process term 'adaptive immune response' between patients with COVID-19 or influenza, a significant increase of these 83 immunoglobulin genes was detected in patients with COVID-19 compared to patients with influenza.*

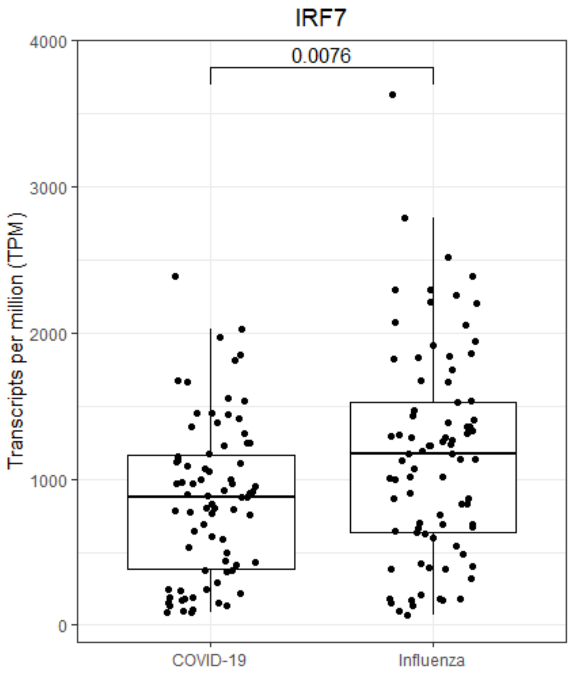


***Supplementary figure 5: Significant difference in IRF7 abundancy between patients with COVID-19 or influenza.*** *The mean abundancy in patients with influenza is 1162 transcripts per million (TPM). This is significantly lower in patients with COVID-19 at 861 TPM (p-value $7.60 \cdot 10^{-03}$). Statistical testing done with Wilcoxon test.*

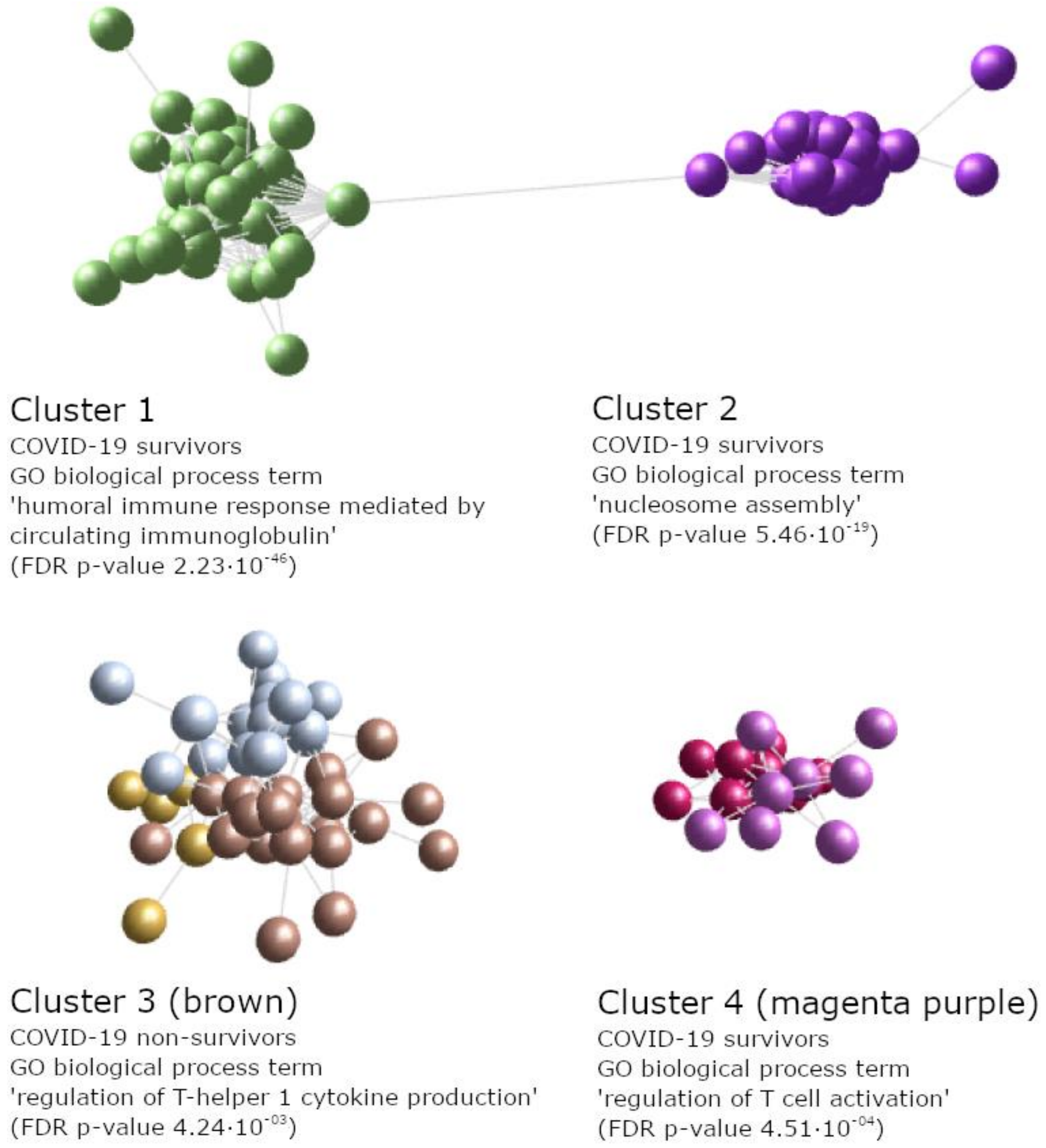


***Supplementary figure 6: Top 4 clusters identified between COVID-19 survivors and non-survivors.*** *For each cluster the predicted Gene Ontology (GO) biological process term identified by ToppGene are given, together with enrichment found in either COVID-19 survivors or non-survivors. From these top 4 clusters combinations of 100 genes were selected as potential predictor variables of COVID-19 outcome using Boosted Logistic Regression, Bayesian Generalised Linear and RandomForest models.*
