## Additional file 5: Differential splicing results and discussion for "Distinct immune responses in patients infected with influenza or SARS-CoV-2, and in COVID-19 survivors, characterised by transcriptomic and cellular abundance differences in blood"

#### Assessment of differential splicing between patient groups

Differential splicing was surveyed using three different methods between patients with COVID-19 versus patients with influenza, and between COVID-19 patients that survived versus those that died. The three tools utilised gave markedly different results. rMATs did not identify any significant differential splicing events between the patient groups. When comparing patients with COVID-19 or influenza, LeafCutter detected 235 differential splicing events with effect size (delta percent-spliced-in (PSI)) >10% in 97 different genes (**Additional file 4**). In the overlapping set of 9,794 genes assessed by both LeafCutter and EdgeR, four genes *(SRGAP2, SRGAP2B, ZBTB16* and *IGHG1*) were both differentially spliced and differentially expressed (Fisher's exact p=0.006). Between COVID-19 survivors and non-survivors, LeafCutter found 95 significant differential splicing events in 47 different genes (**Additional file 4**). Here, 8,527 genes were assessed by both LeafCutter and EdgeR, and four genes (*AC012368.1, IGHG1, MMP8* and *IGHG3*) were found to be both differentially spliced and differentially expressed (Fisher's Exact p=0.009). MAJIQ found a single differential splicing event between patients with COVID-19 or influenza in the gene *FRMD3*, and five differential splicing events in four genes (*CPNE5, IGHG3, PRKXP1 and ANKRD36*) between patients that survived COVID-19 and those that died (**Additional file 4**). Alternative splicing events in *FRMD3*, *CPNE5* and *IGHG3* were detected by both LeafCutter and MAJIQ.

### Discussion

Differential splicing analysis conducted with rMATs, LeafCutter and MAJIQ revealed differentially spliced genes between patients with COVID-19 or influenza, and between patients that survived COVID-19 and those that did not. The differentially spliced genes did not seem to cluster into pathways or processes. Although differentially spliced genes did overlap significantly with differentially expressed genes, most genes displaying differential splicing were not differentially expressed, suggesting independent mechanisms govern the differences in expression and splicing observed between groups. As has been previously reported (1), we found a high degree of variability in the number of differential splicing events detected by the three tools utilised, although the majority of events detected by MAJIQ were also found by LeafCutter. This supports the use of multiple differential splicing tools to maximise detection of events, and highlights the need for further development and optimisation of methodology for differential splicing detection.

#### References


1. Mehmood A, Laiho A, Venäläinen MS, McGlinchey AJ, Wang N, Elo LL. Systematic evaluation of differential splicing tools for RNA-seq studies. Brief Bioinform. 2020 Dec 1;21(6):2052–65.
